# Decoding the hallmarks of GLP-1RA weight-loss super responders

**DOI:** 10.1101/2025.11.15.25340314

**Authors:** AJ Venkatakrishnan, Karthik Murugadoss, Venky Soundararajan

## Abstract

Glucagon-like peptide-1 receptor agonists (GLP-1RAs) have reshaped obesity treatment, yet weight-loss outcomes remain highly uneven in real-world care. Using a federated biomedical platform integrating 23 million de-identified U.S. patient records, we analyzed 135,349 individuals treated with GLP-1RAs and stratified them as “super responders” (>15% weight loss), “moderate responders” (5–15% weight loss), “minimal weight-loss group” (<5% weight loss), and “weight regainers”. super responders reversed nearly two decades of age-associated weight gain in one year, representing approximately a decade more weight reversal than moderate responders. Compared with Wegovy (semaglutide), Zepbound (tirzepatide) showed 47% higher odds (CI: 33–61%) of super-response and 30% lower odds (CI: 23–37%) of minimal weight-loss. Likewise, relative to Ozempic (semaglutide), Mounjaro (tirzepatide) showed 284% (CI: 265–304%) higher odds of super-response and 48% (CI: 46–51%) lower odds of minimal weight-loss. AI-enabled curation processed more than 14 million clinical notes and 15 million structured records covering 1,426 disease terms across the year before and after GLP-1RA initiation. Wegovy and Ozempic super responders showed marked post-treatment increases in vomiting compared with pre-treatment baselines, as reflected by pre-to-post rate ratios (RR 0.37, p=0.014 and RR 0.09, p<0.001). In contrast, Zepbound super responders showed significantly lower post-treatment vomiting relative to baseline (RR 2.34, p<0.001), indicating brand-specific gastrointestinal tolerability profiles. Ozempic (RR 0.24, p<0.001) and Mounjaro (RR 0.17, p<0.001) super responders each showed significant post-treatment increases in diagnoses of protein–energy malnutrition, suggesting a need for whole-body compositional imaging to distinguish beneficial fat loss from unintended lean-mass loss. Novel signals for therapeutic expansion also emerged. Compared with pre-treatment baselines, Zepbound showed significantly reduced post-treatment encounters for recurrent major depressive disorder (pre-to-post RR 12.6, p<0.001) and asthma (pre-to-post RR 2.6, p<0.001). Patient stratification prior to therapy initiation revealed pre-treatment signatures that can guide GLP-1RA choice, with Zepbound super responders showing lower sleep apnea prevalence (baseline RR 0.42, p<0.001) and higher muscle stiffness prevalence (baseline RR 2.4, p=0.037). This study pinpoints actionable physiological signatures and GLP-1RA brand-specific opportunities that emerge from heterogeneous real-world responses, outlining a map for guided precision obesity interventions.

## Introduction

Obesity is a rapidly escalating global public health emergency projected to affect more than 1.1 billion adults by 2030^1^, placing growing strain on health systems worldwide. It fuels rising epidemics of diabetes, cardiovascular conditions, liver disease, and cancer, collectively responsible for more than 1.6 million premature deaths worldwide each year^1^. Despite extensive public health interventions, conventional approaches such as lifestyle modification and bariatric surgery continue to show limited and inconsistent long-term results.

The advent of glucagon-like peptide-1 receptor agonists (GLP-1RAs), originally developed for type 2 diabetes, has transformed obesity treatment^2^. From early agents such as exenatide and liraglutide to next-generation analogs including semaglutide and tirzepatide, this therapeutic class now spans multiple branded formulations with distinct dose ranges and administration regimens (Table S1). GLP-1RAs act through central pathways that suppress appetite and enhance satiety^3^, and through peripheral pathways that improve insulin secretion and glycemic control^4^. Their substantial and durable weight-loss effects, combined with cardiovascular and metabolic benefits, have positioned GLP-1RAs as a cornerstone of modern obesity care^5,6^. Nevertheless, individual weight-loss responses vary widely^7^, underscoring an urgent need for precision therapeutics. Clinical trials and real-world studies report mean weight reductions of 15–20% in patients with obesity or type 2 diabetes mellitus^8^, together with improvements in cardiovascular risk^9^, yet these averages mask profound inter-individual variability. Prior work has been constrained by limited longitudinal follow-up and has not defined clinically distinct responder phenotypes.^9,10^ Addressing this variability is essential to move beyond uniform treatment paradigms and to identify biological and behavioral determinants of exceptional response as well as the drivers of broader heterogeneity in weight-loss outcomes^11^.

Recent advances in electronic health records (EHRs) and AI-assisted curation of unstructured longitudinal data now enable population-level modeling of treatment trajectories.^12–14^ Here, we describe a federated biomedical knowledge platform integrating 23 million de-identified U.S. patient records to model longitudinal physiology, behavior, and therapeutic response among 135,349 individuals prescribed a GLP-1RA (**Figure 1a**). To our knowledge, this represents one of the largest real-world multimodal cohorts assembled to investigate heterogeneity in GLP-1RA treatment responses. Within this cohort, we defined four distinct weight-loss trajectories during the first treatment year: super responders (>15% weight loss), moderate responders (5–15% weight loss), minimal weight-loss group (<5% weight loss), and weight regainers. Using validated AI models for unstructured EHR curation^12,15–18^, we identified pre-treatment phenotypic signatures predictive of GLP-1RA super-response and catalogued brand-specific therapeutic expansion opportunities and adverse-event profiles. These computable clinical phenotypes may illuminate the mechanistic basis of heterogeneity in GLP-1RA effectiveness and inform the design of next-generation precision obesity therapeutics.

**Figure 1.**
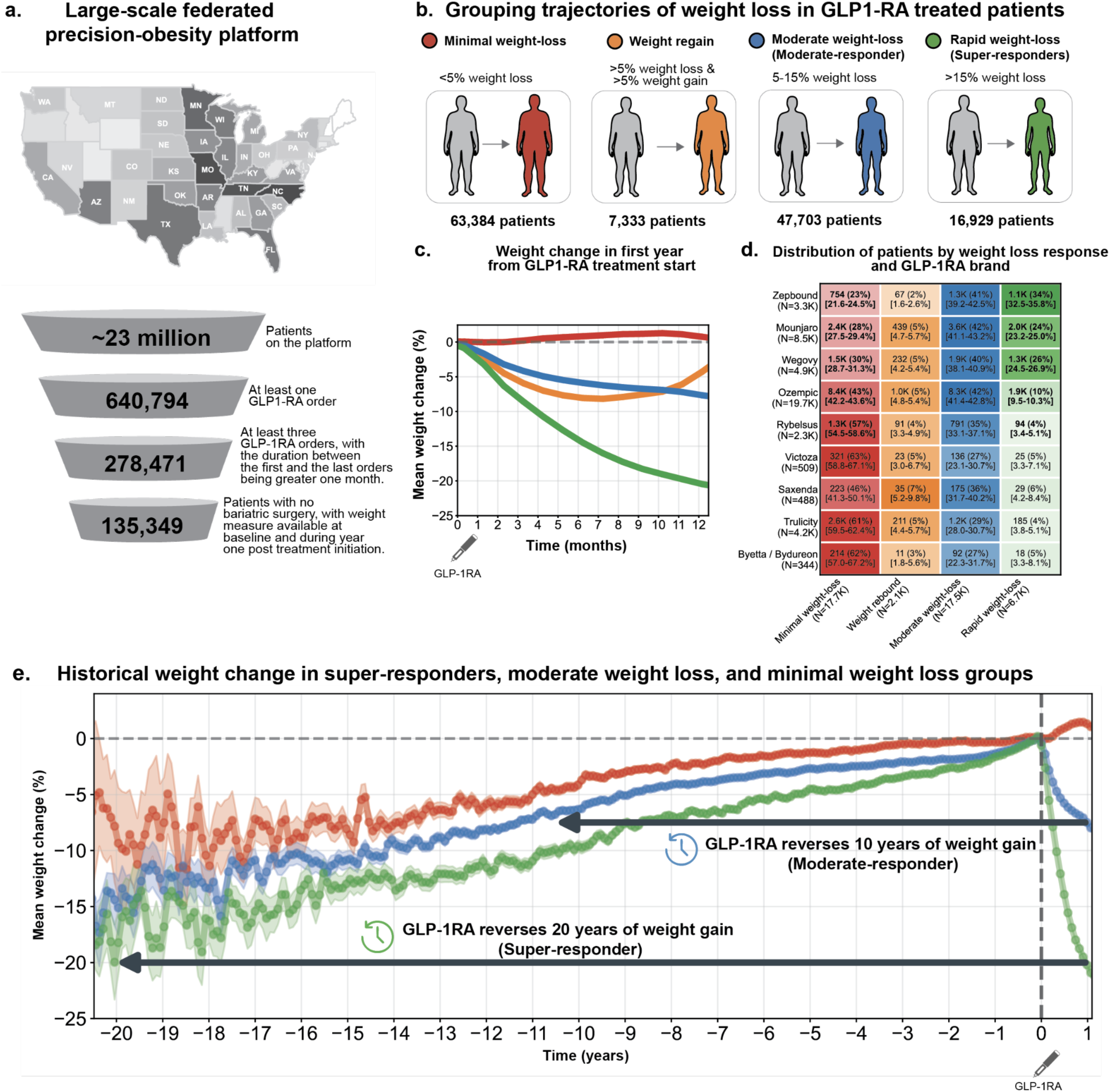
| Defining GLP-1RA treatment trajectories using a large-scale precision obesity platform. **(a.)** Overview of the federated precision obesity platform comprising approximately 23 million de-identified patient records. Of these, 640,794 patients had at least one GLP-1RA order, 278,471 met exposure criteria (>=3 prescriptions spanning >=1 month), and 135,349 had weight measurements available before and after treatment with no history of bariatric surgery, forming the final analytic cohort. (**b.**) Weight-loss trajectory classification in GLP-1RA-treated patients. Individuals were grouped into four categories based on weight change during the first year of GLP-1RA treatment: *minimal weight-loss group* (<5%; red), *weight regain* (>5% loss followed by >5% gain; orange), *moderate responders* (5–15% weight loss; blue), and *super responders* (>15% weight loss, green). **(c.)** Mean first-year post-treatment weight trajectories for the four response categories. **(d.)** Distribution of GLP-1RA brands across the four response categories. Heatmap shows the number and percentage of patients (with 95% CI) in each category for semaglutide (Ozempic, Wegovy) and tirzepatide (Mounjaro, Zepbound) formulations, highlighting substantial brand-specific differences in response profiles. **(e.)** Historical weight trajectories reveal long-term reversal of weight gain among GLP-1RA responders. super responders reversed approximately 20 years of progressive weight gain within one year of therapy, moderate responders reversed roughly 10 years, while the minimal weight-loss group showed little net reversal. Shaded regions represent 95% confidence intervals.

## Results

### Heterogeneous GLP-1RA weight-loss trajectories and reversal of long-term weight gain patterns

We stratified 135,349 GLP-1RA-treated patients into four weight-loss response categories (**Figure 1b**): minimal weight-loss (<5 percent, N=63,384), weight regain (>5 percent gain, N=7,333), moderate weight-loss (5–15 percent, N=47,703), and rapid weight-loss or “super responders” (>15 percent, N=16,929). Mean weight trajectories diverged within the first three months of treatment, after which the minimal weight-loss group remained weight-stable, moderate responders reached a shallow plateau, super responders continued to lose weight throughout the year, and the weight regain group showed a reversal of early weight loss (**Figure 1c**). super responders achieved an average reduction of approximately 18 percent of baseline body weight by one year, compared with roughly 10 percent in moderate responders and negligible change in the minimal weight-loss group. These trajectories highlight pronounced interindividual variability in real-world GLP-1RA effectiveness and reveal distinct physiological response patterns that emerge early in treatment.

Response distributions differed markedly across GLP-1RA formulations and branded products (**Figure 1d**, Table S2). Zepbound (1,118 super responders, 754 minimal weight-loss group) showed significantly higher odds of super-response (OR 1.47, 95 percent CI 1.33–1.61, p<0.001) and lower odds of minimal weight-loss (OR 0.70, 95 percent CI 0.63–0.77, p<0.001) relative to Wegovy (1,258 super responders, 1,468 minimal weight-loss group). Mounjaro (2,039 super responders, 2,410 minimal weight-loss group) showed markedly higher odds of super-response (OR 2.84, 95 percent CI 2.65–3.04, p<0.001) and lower odds of minimal weight-loss (OR 0.52, 95 percent CI 0.49–0.54, p<0.001) compared with Ozempic (1,941 super responders, 8,436 minimal weight-loss group). Moderate-response rates remained consistent across brands, ranging from 40 to 42 percent (41 percent for Zepbound, 42 percent for Mounjaro, 40 percent for Wegovy, and 42 percent for Ozempic), indicating that moderate weight loss was the predominant response phenotype. Earlier GLP-1RAs such as liraglutide (Saxenda) and dulaglutide (Trulicity) were dominated by the minimal weight-loss group in the real-world setting (**Figure 1d)**.

super responders compared to moderate responders and the minimal weight-loss group, were modestly younger (mean age approximately 51 years versus 55 years respectively) and predominantly female (approximately 80 percent versus 58–65 percent respectively), with the highest female representation observed in Zepbound (82.5 percent) and Wegovy (88.3 percent) cohorts (Table S2) in super responders. Across response groups, approximately 2 percent of patients were pregnant in the year prior to GLP-1RA initiation, a factor that may influence baseline weight trajectories. Racial and ethnic distributions differed across response categories, with Caucasian patients enriched among super responders compared to minimal weight-loss patients (90 percent versus 80 percent respectively), whereas lower representation of African American (2.2 percent versus 9.1 percent) and Hispanic patients (1.1 percent versus 5.1 percent) were correspondingly depleted among super responders compared with minimal weight-loss patients (Table S2). Baseline body mass index (BMI) was similar across response groups at approximately 36 kg/m². Type 2 diabetes prevalence was similar between super responders and the minimal weight-loss group for Mounjaro (39.8 versus 39.9 percent) and Ozempic (49.3 versus 52.0 percent), whereas Zepbound (<1 percent) and Wegovy (<2 percent) cohorts showed substantially lower diabetes prevalence across all response groups, consistent with their predominant use in obesity without diabetes (Table S3). Together, these demographic and clinical patterns reveal distinct response-enriched patient segments across GLP-1RA brands and underscore the opportunity for more individualized therapeutic selection.

It is important to note that observed differences in treatment outcomes may partly reflect variability in treatment exposure, including patient’s compliance, duration of prescription and prescription frequency (Figure S1, Table S4). Ozempic (approved December 2017) users had median prescription durations of approximately 14 to 15 months with 8 to 9 prescriptions per patient across response categories. Wegovy (approved June 2021) showed median prescription durations of roughly 8 to 12 months with 6 to 11 prescriptions per patient. Tirzepatide-based therapies Mounjaro (approved May 2022) and Zepbound (approved November 2023) had shorter median prescription durations of about 6 to 12 months, with 6 to 14 prescriptions per patient. These disparities likely reflect differences in time on market, indicated populations, clinical availability, refill logistics, insurance coverage, and titration schedules, all of which may influence real-world weight-loss durability. Clinical engagement, measured by monthly documentation frequency, remained similar across GLP-1RAs and response groups before and after treatment, at a median of 2 to 4 clinical documents per patient per month (Table S4).

Longitudinal reconstruction of pre-treatment body-weight trajectories revealed that GLP-1RA therapy substantially reversed decades of gradual weight gain (**Figure 1e**). On average, super responders experienced a reversal of approximately 20 years of accumulated weight gain, whereas moderate responders reversed roughly 10 years. In contrast, the minimal weight-loss group showed near-flat pre-treatment trajectories and minimal net reversal after GLP-1RA initiation during the first treatment year. A modest late-year weight decline was observed, warranting continued monitoring of the minimal weight-loss group into the second treatment year, particularly for Mounjaro, Zepbound, and Wegovy (Figure S2-S3).

### Post-treatment disease profiles reveal differential adverse events and brand-specific therapeutic opportunities across GLP-1RA response groups

We next investigated how pre– and post-treatment disease prevalence patterns differed across stratified weight-loss response groups for patients treated with Zepbound, Mounjaro, Wegovy, and Ozempic using rate ratios derived from one year before and one year after treatment initiation. A total of 13.9 million unstructured clinical notes and 15 million structured diagnosis records were curated using validated language models to extract 1,426 unique disease terms (see *Methods*; **Figure 2a**).

**Figure 2.**
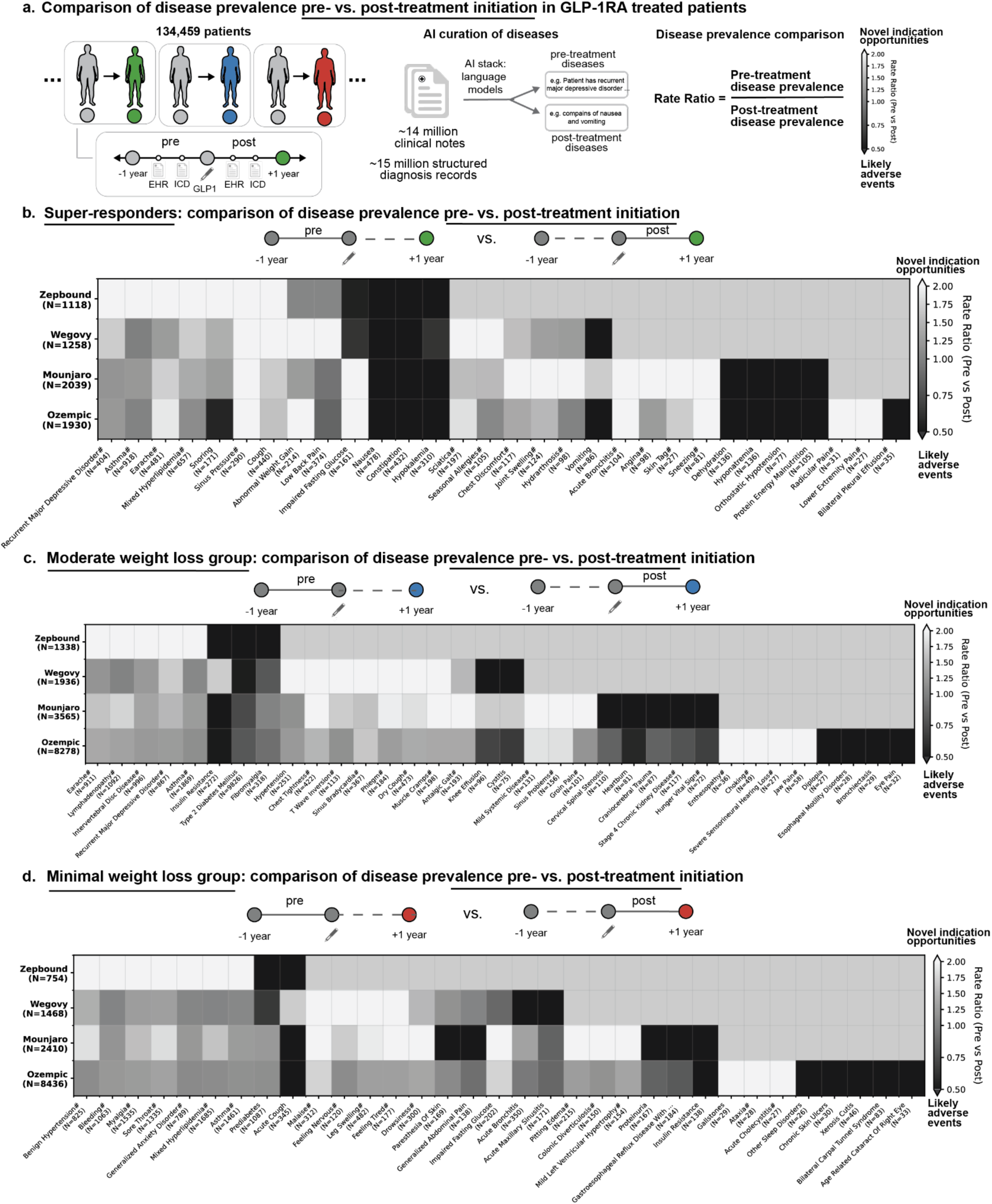
| Comparison of disease prevalence before and after GLP-1RA treatment initiation across weight-loss response groups. **(a.)** Schematic overview of the disease prevalence analysis in 134,459 GLP-1RA–treated patients. Longitudinal electronic health record (EHR) and ICD data were paired with natural language–processed clinical notes (approximately 14 million clinical notes) to identify diseases enriched before or after treatment initiation (diseases labeled with # indicate text-derived diagnoses). Pre– and post-treatment windows spanned one year before and after the first GLP-1RA prescription. The ratio of pre-treatment to post-treatment disease prevalence (Rate Ratio, RR) was used to distinguish conditions that decrease after treatment (RR>1; likely treatment benefits; white boxes in heatmap) versus those that increase (RR<1; potential side effects; black boxes in heatmap). Row N values represent total cohort size for each GLP-1RA brand, while column N values represent total pre-treatment prevalence count for each disease across all four GLP-1RA brands **(b.)** *Super responders:* Heatmap of pre-versus post-treatment disease prevalence rate ratios (pre/post) for the super responder group across four GLP-1RA brands (Zepbound, Wegovy, Mounjaro, Ozempic). **(c.)** *Moderate responders:* Heatmap of pre-versus post-treatment disease prevalence rate ratios (pre/post) for the moderate responder group across four GLP-1RA brands. **(d.)** *Minimal weight-loss group:* Heatmap of pre-versus post-treatment disease prevalence rate ratios (pre/post) for the minimal weight-loss group across four GLP-1RA brands.

We identified 36 potential treatment-associated adverse events in at least one brand (p<0.001, RR≤0.5 indicating higher post-treatment prevalence), encompassing both shared and brand-specific patterns (Table S5). Key findings included shared gastrointestinal and metabolic complications across brands, together with drug-specific differences in tolerability that were consistent with the direction and magnitude of the heatmap rate ratios shown for super responders (**Figure 2b**), moderate responders (**Figure 2c**), and the minimal weight-loss group (**Figure 2d**). super responders across Zepbound, Mounjaro, Wegovy, and Ozempic exhibited consistent post-treatment increases in gastrointestinal and metabolic complications, reflected by RR values less than 1, which indicate higher post-treatment prevalence. These included nausea (RR 0.35–0.43, p<0.001), constipation (RR 0.27–0.39, p<0.001), and hypokalemia (RR 0.18–0.64, p<0.001 except for Wegovy).

Vomiting demonstrated divergent patterns between tirzepatide and semaglutide in super responders (Table S5). Zepbound users showed significant post-treatment reductions (RR 2.34, p<0.001), Mounjaro super responders exhibited no significant change (RR 1.07, p=0.603), whereas semaglutide users showed substantial post-treatment increases (Wegovy RR 0.37, p=0.014; Ozempic RR 0.09, p<0.001). Diarrhea prevalence decreased consistently with tirzepatide formulations but not with semaglutide formulations (Table S5). Zepbound users showed reductions across all response groups (RR 1.9–2.5, p<0.001), and Mounjaro users showed reductions in minimal and moderate responders (RR 1.2–1.5, p<0.001) with a similar trend in super responders that did not reach statistical significance (RR 1.2, p=0.12). Semaglutide formulations showed minimal overall change (RR 0.7–1.3).

Prevalence of protein–energy malnutrition diagnosis showed a weight-loss–dependent pattern for select GLP-1RAs: prevalence increased post-treatment in Ozempic and Mounjaro super responders (RR 0.24 and 0.17, both p<0.001) and in Ozempic moderate responders (RR 0.6, p=0.037), while Ozempic the minimal weight-loss group showed decreased prevalence (RR 2.5, p=0.001). This pattern was not observed in Zepbound or Wegovy users, suggesting drug-specific differences in appetite suppression intensity and nutritional adequacy maintenance despite comparable weight-loss outcomes (Table S5).

Remarkably, GLP-1RA treatment was also associated with broad reductions in airway– and upper-respiratory–related symptoms across several brands, with the most pronounced effects observed in Zepbound users (**Figure 2b**). For example, across all treatment groups, the prevalence of sinus pressure (RR=2.5-6.4, p<0.005) and rhinorrhea (RR=1.3-8.0, p<0.05) declined consistently following treatment initiation. Among Zepbound (tirzepatide) users, the reduction of airway-related symptoms was particularly pronounced: snoring decreased in super responders (RR 8.4, p<0.001), moderate responders (RR 4.2, p<0.001), and the minimal weight-loss group (RR 7.2, p<0.001), paralleled by marked declines in obstructive sleep apnea (RR 2.76-3.43, p<0.001) and asthma (RR 2.62-3.59, p<0.001). Mounjaro (tirzepatide) also showed reduced prevalence of obstructive sleep apnea in the moderate responders and minimal weight-loss group (RR>1.3, p<0.001) while semaglutide formulations demonstrated no appreciable change (RR 0.85-1.1). Likewise, reduction in asthma prevalence was observed upon initiating Mounjaro across all weight-loss groups (RR>1.2, p<0.05), but no significant reduction in asthma was observed with either of the semaglutide formulations (Wegovy and Ozempic RR 0.8-1.1).

The prevalence of earache declined substantially across Zepbound cohorts, most notably in moderate responders (RR 13.4, p<0.001). Similarly, post-treatment earache prevalence also declined in Mounjaro users (RR 1.35-1.91, p<0.001) and Ozempic users (RR 1.62-1.76, p<0.001). Mounjaro super responders also exhibited large post-treatment reductions in acute bronchitis (RR 8.8, p<0.001). These ear, nasal, and respiratory symptoms reductions exhibit some of the highest RR values across the entire matrix (**Figure 2b-c**).

Further, Zepbound users exhibited striking reductions in recurrent major depressive disorder across all response categories (super responders RR 12.6, moderate responders RR 9.6, minimal weight-loss group RR 8.0; all p<0.001), a pattern not observed in Wegovy, Mounjaro, or Ozempic users (RR 0.8–1.4), suggesting a potential Zepbound-specific depression benefit that warrants further investigation (**Figure 2b-d**).

Collectively, these findings reveal substantial post-treatment reductions in sinus pressure, rhinorrhea, snoring, obstructive sleep apnea, asthma, earache, acute bronchitis, and recurrent major depressive disorder in real-world settings, with the most pronounced changes observed among tirzepatide users, particularly Zepbound. These effects were not uniform across GLP-1RA formulations and were limited to the specific symptoms and diagnoses identified in our analysis of 1,426 disease terms. The observed patterns should therefore be interpreted strictly as condition-specific signals, rather than evidence for broader category-level improvements, and they motivate targeted mechanistic and clinical follow-up to understand why these particular symptoms and diagnoses change after GLP-1RA initiation.

### Pre-treatment disease signatures reveal brand-specific physiological entry points to GLP-1RA response trajectories

To investigate whether differential pre-treatment health profiles predispose patients to divergent GLP-1RA outcomes, we performed propensity-matched comparisons of pre-treatment disease prevalence between: (1) super responders versus moderate responders and (2) super responders versus minimal weight-loss patients. The cohort pairs were propensity matched exclusively on age and sex (see *Methods*). Cohort pairs demonstrated comparable baseline characteristics, including pregnancy rates (approximately 2 percent, Table S6), BMI (approximately 36 kg/m², Table S7), nearly identical type 2 diabetes prevalence (within approximately two percentage points, Table S7), and similar clinical engagement (median three clinical documents per patient per month, Table S8). Brand-specific pre-treatment signatures emerged that differentiated super responders from both minimal and moderate responders (**Figure 3a-b**).

**Figure 3.**
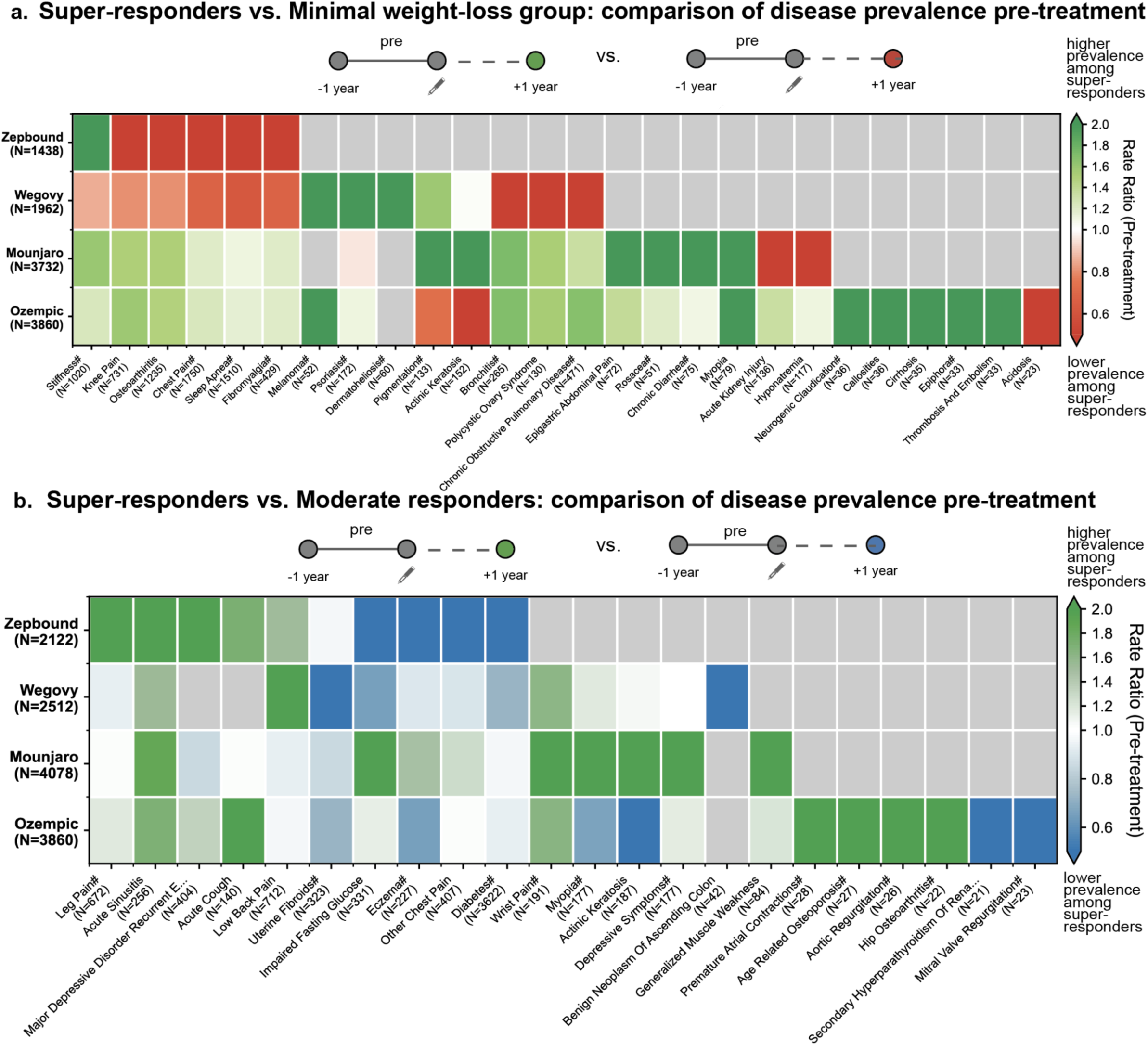
| Pre-treatment disease signatures distinguishing GLP-1RA super responders from moderate and minimal weight-loss groups. **(a.)** Heatmap comparing pre-treatment disease prevalence between super responders and the minimal weight-loss group across four GLP-1RA brands (Zepbound, Wegovy, Mounjaro, Ozempic). Each cell displays the pre-treatment Rate Ratio (RR), calculated as the underlying prevalence in super responders divided by the underlying prevalence in the minimal weight-loss group in the one year prior to GLP-1RA treatment initiation. Green shading indicates RR > 1 and red shading indicates RR < 1. Row N values represent total cohort size for each propensity-matched comparison (matched pairs × 2), and column N values represent the total pre-treatment prevalence count for each disease across all four brands. Diseases labeled with “#” denote text-derived diagnoses identified from AI-curated clinical notes. **(b.)** Heatmap comparing pre-treatment disease prevalence between super responders and the moderate responders across four GLP-1RA brands. Each cell displays the pre-treatment Rate Ratio (RR), calculated as the underlying prevalence in super responders divided by the underlying prevalence in the moderate responders in the one year prior to GLP-1RA treatment initiation. Green shading indicates RR > 1 and blue shading indicates RR < 1. Row N values represent total cohort size for each propensity-matched comparison (matched pairs × 2), and column N values represent the total pre-treatment prevalence count for each disease across all four brands. Diseases labeled with “#” denote text-derived diagnoses identified from AI-curated clinical notes.

### Zepbound super responders

Compared with the Zepbound minimal weight-loss cohort (N=719 matched pairs), Zepbound super responders had higher pre-treatment prevalence of muscle stiffness (RR 2.4, p=0.037) and lower prevalence of fibromyalgia (RR 0.2, p=0.002), knee pain (RR 0.5, p=0.014), chest pain (RR 0.4, p<0.001), osteoarthritis (RR 0.5, p=0.001), and obstructive sleep apnea (RR 0.4, p<0.001; **Figure 3a**).

In a separate propensity-matched comparison versus Zepbound moderate responders (N=1,061 matched pairs), Zepbound super responders showed higher pre-treatment prevalence of acute sinusitis (RR 3.1, p=0.031), recurrent major depressive disorder (RR 2.1, p=0.042), and leg pain (RR 3.7, p=0.003) (**Figure 3b**), and lower prevalence of eczema (RR 0.2, p=0.008), impaired fasting glycemia (RR 0.3, p=0.027), and diabetes (RR 0.2, p=0.002).

### Wegovy super responders

Wegovy super responders had significantly higher pre-treatment prevalence relative to the Wegovy minimal weight-loss cohort (*N*=981 matched pairs) for psoriasis (RR=2.5, p=0.033), dermatoheliosis (RR=2.0, p=0.044), and melanoma (RR=4.4, p=0.008; **Figure 3a**) and lower pre-treatment prevalence for bronchitis (RR=0.2, p<0.001), chronic obstructive pulmonary disease (RR=0.2, p<0.001), and polycystic ovarian syndrome (RR=0.2, p<0.001). Further, compared with Wegovy moderate responders (*N*=1,256 matched pairs), Wegovy-super responders had higher pre-treatment prevalence for low back pain (RR=5.4, p=0.007), lower prevalence for uterine fibroids (RR=0.4, p=0.012; **Figure 3b**).

### Mounjaro super responders

Mounjaro super responders had significantly higher pre-treatment prevalence relative to the Mounjaro minimal weight-loss cohort (*N*=1,866 matched pairs) for myopia (RR=2.9, p=0.01), chronic diarrhea (RR=5.4, p=0.001), epigastric abdominal pain (RR=5.4, p=0.001), rosacea (RR=5.4, p=0.001), actinic keratosis (RR=7.4, p<0.001), and pigmentation (RR=6.2, p<0.05), and lower prevalence of acute kidney injury (RR=0.34, p=0.010) and hyponatremia (RR=0.37, p=0.029; **Figure 3a**). Further, compared with Mounjaro moderate responders (*N*=2,039 matched pairs), Mounjaro super responders were noted for generalized muscle weakness (RR=4.4, p=0.033), depressive symptoms (RR=5.4, p=0.007), actinic keratosis (RR=2.8, p=0.021), myopia (RR=3.1, p=0.031), wrist pain (RR=3.0, p=0.007), and impaired fasting glucose (RR=2.7, p=0.007; **Figure 3b**).

### Ozempic super responders

Ozempic super responders had significantly higher pre-treatment prevalence relative to Ozempic minimal weight-loss patients (*N*=1,930 matched pairs) for myopia (RR 7.2, p<0.001), neurogenic claudication (RR 7.2, p<0.001), callosities (RR 7.2, p<0.001), cirrhosis (RR 7.0, p<0.001), epiphora (RR 6.6, p<0.001), and embolism (RR 6.6, p<0.001), and lower pre-treatment prevalence of acidosis (RR 0.2, p=0.006) and actinic keratosis (RR 0.3, p=0.002; **Figure 3a**). Further, comparing with Ozempic moderate responders (*N*=1,930 matched pairs), Ozempic super responders exhibited higher relative prevalence of acute cough (RR 5.2, p=0.008), premature atrial contractions (RR 5.6, p=0.005), age-related osteoporosis (RR 5.4, p<0.001), aortic regurgitation (RR 5.2, p=0.008), hip osteoarthritis (RR 4.4, p=0.033), and lower pre-treatment prevalence of actinic keratosis (RR 0.3, p=0.002), secondary hyperparathyroidism (RR 0.2, p=0.044), hypotension (RR 0.2, p=0.023) and mitral valve regurgitation (RR 0.2, p=0.023; **Figure 3b**).

Taken together, these pre-treatment signatures reveal brand-specific physiological entry points into GLP-1RA response trajectories and suggest that distinct baseline phenotypes may predispose certain patients to exceptional weight-loss outcomes.

## Discussion

### Decades-long weight-gain reversal in a subset of GLP-1RA–treated patients suggest a framework to study “metabolic age”

In the new era of incretin-based obesity pharmacotherapy, understanding and leveraging the marked heterogeneity in individual responses is critical for optimizing therapeutic selection. The inter-individual variation is striking, with outcomes ranging from weight regain and minimal weight loss on significant patient populations to equally significant super responders who benefit from more than 15 percent weight loss on some of the same agents. Reconstructing the longitudinal weight history journeys enabled a striking finding that super responders erase more than twenty years of accumulated weight gain within a single year of therapy, whereas the larger population of moderate responders still reverse approximately ten years of their prior weight gain trajectory **(Figure 1e)**. These decade-level weight reversals illustrate the profound benefit achieved by some patients and emphasize how unevenly individuals respond to the same therapy. In the context of recent discussion that GLP-1RAs may modulate aging-related physiology^19^, this wide response spectrum highlights that any potential healthspan benefits are unlikely to be uniform and instead depend heavily on individual biological context. In parallel with advances that compute organ ages such as cardiac age^20^ and brain age^21^, our findings motivate an inquiry into the existence and characterization of “metabolic age”. Further our study suggests that precision stratification, not expanded access alone, will determine whether incretin therapies achieve population-level impact.

### Specific respiratory, musculoskeletal, and dermatologic signatures associated with super responder baseline profiles and post-treatment patterns

The clinical signatures decoded by this study reveal consistent brand-associated response patterns with specific baseline phenotypes and post-treatment changes aligning with emerging knowledge of GLP-1RA biology and multi-system effects (**Figure 4**)^19^. Following treatment initiation, tirzepatide demonstrated marked pre-to-post reductions in obstructive sleep apnea across both Zepbound and Mounjaro cohorts, while semaglutide formulations showed no appreciable change (**Figure 2b**). Indeed, these real-world respiratory signals are directionally aligned with SURMOUNT-OSA phase 3 trial results^22^, in which tirzepatide reduced the apnea–hypopnea index in individuals with obesity and obstructive sleep apnea. Interestingly, in our study, Zepbound super responders showed lower baseline sleep apnea prevalence compared to patients who achieved minimal or moderate weight loss (**Figure 4a**). Our findings and the clinical trial results motivate future research into the relationship between GLP-1 biology and sleep apnea.

**Figure 4.**
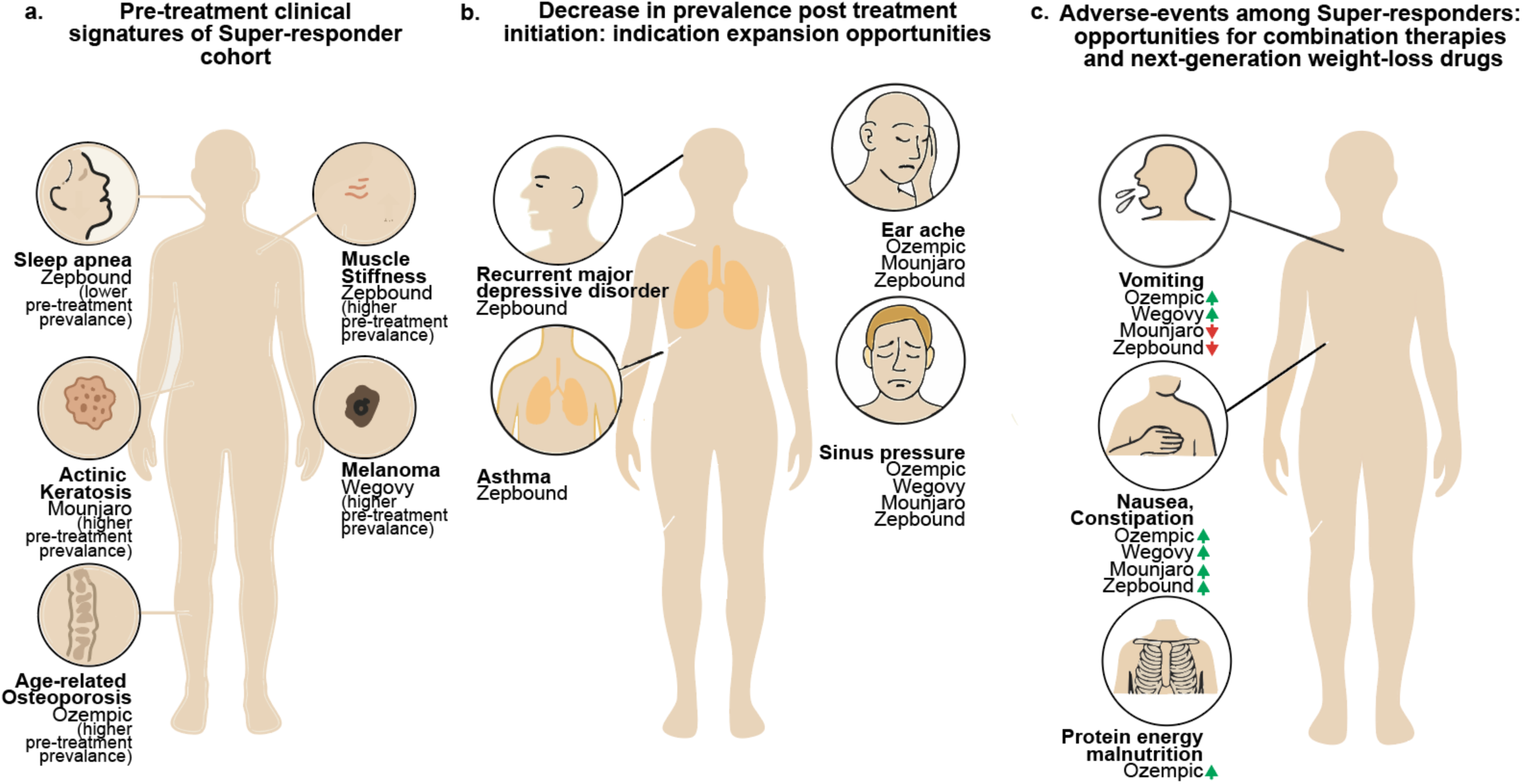
| Clinical signatures and treatment-associated patterns among GLP-1RA super responders. **(a.)** Pre-treatment clinical signatures enriched or depleted in super responder cohorts across Zepbound, Mounjaro, Wegovy, and Ozempic. **(b.)** Post-treatment decreases in disease prevalence among super responders, suggesting potential domains for GLP-1RA indication expansion. **(c.)** Adverse-event patterns among super responders highlighting opportunities for supportive combination therapies and design of future weight-loss agents.

Zepbound super responders showed higher baseline prevalence of muscle stiffness alongside reduced burden of knee pain and osteoarthritis (**Figure 4a**, **Figure 3a**), suggesting that patients with obesity-related muscular dysfunction but preserved joint health may be particularly likely to achieve exceptional weight-loss outcomes with tirzepatide. This is clinically relevant in light of growing interest in muscle-preserving strategies during GLP-1RA-induced weight loss, including trials combining incretin therapies with agents that target muscle mass and function^23,24^. Likewise, the enrichment of age-related osteoporosis in Ozempic super responders points to a subgroup with pre-existing skeletal fragility for whom rapid weight loss may warrant closer bone-health and falls monitoring (**Figure 4a**, **Figure 3b**), consistent with recent analyses showing that semaglutide-associated weight loss can be accompanied by significant reductions in lean mass and bone density in susceptible individuals^25–27^.

The dermatologic signatures in this study similarly map onto emerging data on skin–metabolism crosstalk^28–31^. Mounjaro super responders were enriched for pre-treatment actinic keratosis, and Wegovy super responders for dermatoheliosis and melanoma, indicating that some highly responsive patients carry a background of photoaging and melanoma, which may arise from ultraviolet exposure, dysplastic nevi, or genetic susceptibility^32^. Experimental and translational studies suggest that GLP-1RAs can modulate dermal fibroblast function, reduce oxidative stress, and influence cutaneous immune responses, supporting the concept that skin is one of several peripheral tissues affected by incretin signaling^33^. Our findings do not imply any relationship with melanoma risk, which has not been consistently increased in large GLP-1RA safety studies, but they do underscore that dermatologic history may help define specific super responder phenotypes under semaglutide or tirzepatide therapy^31^.

### Specific neuropsychiatric and pulmonary improvements associated with tirzepatide highlight potential therapeutic avenues beyond metabolic disease

The reduction in certain non-metabolic conditions among super responders, particularly in Zepbound users, provides hypothesis-generating evidence for indication expansion (**Figure 4b**). In our cohort, tirzepatide super responders had fewer encounters for recurrent major depressive disorder and asthma after treatment initiation. These observations are directionally consistent with emerging literature suggesting that GLP-1RAs can modulate neuroinflammatory and immune pathways relevant to mood and airway disease, including preclinical work showing that GLP-1 receptor activation in microglia and other brain-resident cells attenuates neuroinflammation and alters transcriptomic aging signatures in glial populations^34,35^. Single-cell RNA-sequencing and receptor-mapping studies further demonstrate GLP-1R and GIPR expression in discrete neuronal and glial subsets within brainstem and hypothalamic circuits that regulate stress, autonomic tone, and appetite, providing a cellular framework through which incretin signaling could influence depressive symptoms and related neurobehavioral outcomes^36,37^.

Similarly, the asthma and sinus-pressure improvements we observed with tirzepatide are compatible with systems-level data suggesting that GLP-1RAs broadly modulate inflammatory and vascular pathways (**Figure 4b**)^38^. Proteomic studies in semaglutide-treated patients have identified changes in circulating proteins involved in lipid metabolism, immune signaling, and vascular biology beyond what is explained by weight loss alone, supporting a model in which incretin therapies reshape systemic inflammatory networks that also intersect with airway disease^39,40^. In addition, we observed brand-spanning decreases in prevalence of sinus pressure and, for several agents, earache, which may reflect improvements in upper-airway congestion or mucosal inflammation. While these signals should be interpreted cautiously, they illustrate how large-scale EHR analyses, when viewed alongside single-cell and proteomic studies of GLP-1RA action, can surface unexpected hypotheses of potential benefit that merit targeted mechanistic and clinical follow-up.

### Differences in gastrointestinal tolerability between tirzepatide and semaglutide may reflect dual-agonist versus GLP-1–only mechanisms

Across all four GLP-1RA brands in our cohort, nausea and constipation were more prevalent after treatment than before treatment, indicating that these gastrointestinal effects represent class-wide tolerability features rather than agent-specific outliers. However, strikingly, semaglutide super responders experienced increased prevalence of vomiting, whereas Zepbound super responders experienced decreased prevalence of vomiting relative to their pre-treatment baselines **(Figure 4c)**. One possible explanation for this paradoxical divergence is that semaglutide is a pure GLP-1 receptor agonist, producing strong and sustained activation of GLP-1R pathways in the area postrema and nucleus tractus solitarius, both of which are tightly linked to nausea and emesis circuits^41,42^. In contrast, tirzepatide engages both GIP-R and GLP-1R, and emerging preclinical data suggest that GIP receptor activation may attenuate GLP-1-induced vomiting by counterbalancing anorectic and emetic signaling in brainstem autonomic circuits and by modulating vagal afferent tone^43^. This dual-agonism distinction, together with differences in gastric emptying kinetics, receptor internalization, and central penetration, may help explain why post-treatment vomiting increases with semaglutide but decreases in Zepbound super responders, despite greater overall weight-loss efficacy.

Diagnosis of protein-energy malnutrition increased specifically in Ozempic and Mounjaro super responders, but not in Zepbound or Wegovy users, which aligns with body-composition data indicating that potent GLP-1RAs can reduce lean mass as well as fat mass and raises concern for sarcopenia and nutritional depletion in high-response patients^25,44^. These post-treatment patterns demonstrate that each GLP-1RA in our cohort was associated with a distinct constellation of adverse events and symptom improvements, reinforcing the need to assess safety and potential indication expansion at the individual drug level rather than generalizing across the entire class.

### Toward predictive, personalized incretin and non-incretin metabolic therapy: integrating dosing, lifestyle, and multimodal phenotyping

Despite the findings made here, multiple scientific and translational challenges remain to be addressed that will shape the next phase of incretin and non-incretin metabolic therapeutic development. First, granular dosing trajectories, titration patterns, persistence metrics, and adherence data, which influence real-world effectiveness, remain to be fully characterized (Figure S4)^45,46^. Interruptions, re-initiations, and incomplete dose escalation likely contribute meaningfully to the observed real-world variation in treatment outcomes, and patients often have multiple co-existing conditions treated with diverse therapies beyond GLP-1RA treatments that may exert combined influence on weight trajectories at the individual level^47^. Second, the present analysis focuses on brand-exclusive cohorts to enable clean, within-agent response phenotyping; however, many patients in real-world care transition between incretin therapies over time^48^. Although switching behavior is not examined in the present study, it can be analyzed within our dataset and will be evaluated in follow-up work to characterize sequential therapy patterns and their impact on clinical outcomes. Third, although diet quality, physical activity, and sleep patterns were not incorporated into the present analyses, our multimodal platform captures these quality-of-life measurements through questionnaires, flowsheets, and longitudinal clinical note mentions that can be systematically quantified^49–51^. These behavioral factors will be integrated in future work to characterize how lifestyle dynamics shape metabolic therapeutic response trajectories. Similarly, unmeasured socioeconomic factors including food security and healthcare access may vary across response groups and confound observed associations^52,53^. Fourth, the platform contains radiomics derived from longitudinal imaging, along with extensive laboratory trajectories spanning metabolic, endocrine, inflammatory, and lipidomic domains (Figure S6). Integrating these anatomical and molecular modalities will be essential for deeper phenotyping of the physiological pathways that influence response across both incretin and non-incretin metabolic therapies. Likewise, de-identified pharmacogenomic information and cytokine or immune-mediator panels were not studied in the present analysis, limiting our ability to assess genetic modifiers of incretin signaling or molecular inflammatory states that may stratify patients. Multimodal longitudinal inference at scale can elevate pre-treatment signatures into mechanistic insight and enable predictive, personalized metabolic therapy across the expanding landscape of incretin and non-incretin agents. When multimodal AI synthesizes imaging, laboratory data, and clinical histories spanning several decades of care, a striking possibility emerges — that the metabolic arc of aging may be more malleable than previously imagined.

## Methods

### Data source & selection

This study analyzed de-identified EHR data from a network of tertiary clinical centers tied to academic medical centers in the United States through the nference nSights Analytics Platform^55,56^. nference, in collaboration with academic medical center (AMC) data partners provided the de-identified data for this study. nference has established a secure data environment, hosted by and within each of the AMCs, that house the AMC’s de-identified patient data. The provisioning of and access to this data are governed by an expert determination that satisfies the HIPAA Privacy Rule requirements for the de-identification of protected health information. Each AMC’s de-identified data environment is specifically designed and operated to enable access to and analysis of de-identified data without the need for Institutional Review Board (IRB) oversight, approval, or an exemption confirmation. Given these measures, informed consent and IRB review were not required for this study.

### Cohort eligibility criteria and study timeline

From a longitudinal database of 23 million patients, we identified 640,794 patients with at least one GLP-1RA order. To ensure adequate treatment exposure, we required patients to have at least 3 GLP-1RA orders, with the first and last orders spanning a period of at least one month, yielding 278,471 patients. We then excluded patients with a history of bariatric surgery (including gastric bypass, sleeve gastrectomy, gastric banding, and duodenal switch) and required availability of weight measurements during both a baseline period (defined as no more than 90 days prior to the first GLP-1RA prescription) and during the one-year period after the first prescription. The final analytical cohort comprised 135,349 patients with weight trajectory data during the first year of GLP-1RA therapy (**Figure 1a**).

### Weight Trajectory Analysis

Patients were classified into four response categories based on their maximum weight loss within 12 months post-treatment initiation (Figure 1b): super responders (≥15%), moderate responders (5-15%), minimal weight-loss group (<5%), and weight regainers (initial ≥5% loss followed by a weight regain of at least 5%). For each patient, the index date was defined as the date of first GLP-1RA prescription, and baseline weight was determined as the measurement closest to the index date within a 90-day pre-treatment window. Percentage weight change was calculated relative to this baseline for all measurements. To generate population-level trajectories, weight measurements were aggregated into 30-day time windows (±15 days from each time point), and patient-level mean percentage changes were computed within each window (**Figure 1c**, **Figure 1e**). Patient-level mean values were calculated across patients and were then smoothed using a Savitzky–Golay filter (window length = 5, polynomial order = 3) to reduce noise while preserving temporal trends. Uncertainty was quantified using the standard error of the mean (SEM = σ/√n), where σ represents the standard deviation of patient-level means and n represents the number of patients with measurements in each time window. Shaded regions around trajectory lines represent ±1 SEM.

### GLP-1RA Brand Stratification Analysis

To assess medication-specific effects, patients were stratified into mutually exclusive cohorts based on their GLP-1RA exposure history. “Brand-exclusive” cohorts comprised patients who received only a specific branded formulation (Ozempic, Wegovy, Rybelsus, Mounjaro, Zepbound, Victoza, Saxenda, Trulicity, Byetta, or Bydureon) without switching to any other brand or molecule. Bydureon and Bydureon BCise were combined into a single cohort given their pharmacological equivalence. Patient distribution across response categories for each brand was visualized using heatmaps (**Figure 1d**), with cell color intensity scaled independently within each response category column to highlight relative differences across medications. Each cell displayed the patient count and percentage of the drug/brand cohort, accompanied by 95% confidence intervals calculated using the Wilson score method for binomial proportions. Odds ratios (OR) with 95% confidence intervals were calculated to compare treatment response categories between GLP-1 receptor agonist formulations. Statistical significance was assessed using the Wald test for log-transformed odds ratios. P-values < 0.05 were considered statistically significant.

### Propensity Score Matching

To mitigate confounding comparisons between super responders and moderate responders/the minimal weight-loss group, propensity score matching was performed using age and gender as matching variables. Propensity scores were estimated using logistic regression with a maximum of 1,000 iterations and the Limited-memory Broyden-Fletcher-Goldfarb-Shanno (L-BFGS) optimization algorithm, with treatment status (super responders versus minimal weight-loss group, or super responders versus moderate responders) as the outcome. Gender was encoded as a binary variable (male=1, female=0), and patients with missing values in either matching variable were excluded prior to score estimation. Greedy nearest-neighbor matching without replacement was conducted with a 1:1 matching ratio and a caliper width of 0.1 (maximum absolute difference in propensity scores between matched pairs). For each super responder, the nearest available patient from the minimal weight-loss (or moderate weight-loss) cohort within the caliper was selected, and that patient was removed from the matching pool. super responders who could not be matched within the caliper were excluded from subsequent trajectory analyses. This approach yielded balanced cohorts with minimized baseline age and gender differences between weight loss groups.

### Summary Statistics and Statistical Testing

Summary statistics were generated for the overall cohort as well as for drug-exclusive and brand-exclusive cohorts, with analyses performed separately for propensity-matched and unmatched populations (Tables S2-S4, S6-S8). Demographic distributions including age (at first GLP-1RA prescription), gender and race (White, Black/African American, Hispanic, Other/Unknown) were assessed across all cohorts. Baseline clinical characteristics included pregnancy status, Type 2 diabetes mellitus (T2DM) status, body mass index (BMI) and weight, defined as measurements obtained within 90 days prior to the first prescription. Pregnancy status was defined as the presence of one or more pregnancy-associated diagnoses documented in the 1-year period preceding the first GLP-1 RA prescription. T2DM status was defined as the presence of three or more diabetes diagnoses documented in the 5-year period preceding the first GLP-1 RA prescription. The distribution of weight measurements per patient was characterized across both the pretreatment and one-year post-treatment period. The time interval in months between the first and last GLP-1RA prescription was calculated for each patient, and prescription frequency was quantified as the total number of prescriptions received during follow-up. To assess data completeness, baseline monthly clinical notes per patient (quantified over the year prior to the index date) and post-treatment monthly clinical notes per patient (quantified over the 12 months following the index date) were calculated. Categorical comparisons between response groups including gender distribution, T2DM status, pregnant patient status, and race/ethnicity were evaluated using counts (N) and percentages (%), with chi-square tests used for equality of proportions. Statistical significance for categorical comparisons was assessed using p-values, with p<0.05 considered statistically significant. For continuous variables, means with standard deviations (SD) and medians with interquartile ranges (IQR; 25th and 75th percentiles) were calculated.

### Augmented Curation

We employed a multi-stage information extraction (IE) pipeline to extract clinical phenotypes, such as patient diseases and comorbidities, from unstructured clinical notes. The pipeline’s architecture combined fine-tuned Bidirectional Encoder Representations from Transformers (BERT)^54^ models. The workflow first employs a Named Entity Recognition (NER) model to identify clinical entities, including diseases, symptoms, and medications. Subsequently, a series of sequential qualifier models assesses the clinical context for each extracted entity by determining its subject (patient vs. other), temporality (current, past, or hypothetical), and certainty (confirmed, negated, or suspected). This contextualization framework is essential for disambiguating confirmed, current patient symptoms (e.g., “Patient reports persistent nausea”) from negated findings (e.g., “No vomiting”), suspected conditions (e.g., “Rule out gastroparesis”) or prior diagnoses (e.g. “Had sleep apnea last year”). This IE approach has been validated in prior studies^15^, demonstrating high fidelity in capturing conditions and medications from clinical notes with F1 scores of 0.93 and 0.95, respectively. For instance, compared to structured EHR data alone, the diagnosis extraction component captured significantly more individuals with relevant signs and symptoms, including vomiting (Enrichment Factor [EF]: 22.8, 95% CI: [15.2, 41.1]), weight loss (EF: 5.0, 95% CI: [4.1, 6.6]), nausea (EF: 4.3, 95% CI: [3.1, 6.9]), and edema (EF: 3.1, 95% CI: [2.7, 3.7]).

### Longitudinal and Comparative Analysis of Phenotypes in Weight Loss Cohorts

We performed a comparative analysis of disease prevalence by assessing patient cohorts at different time points and against each other across a total set of 1,426 unique diseases. The analysis utilized both structured diagnoses and unstructured phenotypes. These phenotypes were extracted from all available clinical documentation within the defined pre-treatment (1 year to 30 days prior to index) and post-treatment (30 days to 1-year post-index) periods, restricting the output to confirmed, current patient-attributed diseases. Three primary sets of comparisons were conducted: (1) a longitudinal pre-treatment vs. post-treatment analysis, performed separately for rapid, moderate, and minimal weight-loss cohorts (**Figure 2**); (2) a comparative pre-treatment analysis, comparing super responders to both minimal weight-loss group and moderate responders (**Figure 3**); and (3) a comparative post-treatment analysis, comparing super responders to both the minimal weight-loss group and moderate responders (Figure S5). These comparisons were performed independently for four GLP-1RA medications (Zepbound, Mounjaro, Wegovy, and Ozempic). The prevalence of each disease phenotype was compared by calculating a Rate Ratio (RR) and a chi-square test p-value, corrected for False Discovery Rate (FDR) using the Benjamini-Hochberg (BH) procedure. For heatmap visualization, a subset of features was selected by identifying diseases that showed both statistically significant associations (FDR-corrected p<0.05) and demonstrated strong effect sizes (RR > 2.0 or RR < 0.5) in at least one GLP-1RA medication. To construct the final disease list for each heatmap, a maximum of five diseases with the highest RRs and five diseases with the lowest RRs were selected per drug.

### De-identification and HIPAA compliance certification

Prior to analysis, all EHR data were de-identified under an expert determination consistent with the Health Insurance Portability and Accountability Act (HIPAA) Privacy Rule (45 CFR §164.514(b)(1)). The de-identification methodology employed a multi-layered transformation approach to both structured and unstructured data fields^55,56^. In structured data, direct identifiers including patient names and precise geographic locations were excluded entirely, while indirect identifiers underwent specific transformations: patient identifiers, medical record numbers, and accession numbers were replaced with one-way cryptographic hashes using confidential salts to preserve linkage across patient encounters; all dates were shifted backward by patient-specific random offsets (1–31 days) to preserve temporal relationships while obscuring exact event timing; the ZIP codes were truncated to two-digit state-level resolution; and continuous variables including age, height, weight, and body mass index were thresholded to prevent identification of extreme values (for example, ages ≥89 years transformed to ‘89+’ and BMI >40 transformed to ‘40+’). In unstructured clinical text, an ensemble de-identification system that combines attention-based deep learning models with rule-based methods achieved an estimated >99% recall for personally identifiable information (PII) detection, with detected identifiers replaced by plausible fictional surrogates^55^.

### Data Harmonization

To address heterogeneity in EHR data, we harmonized clinical variables including medications, anthropometric measurements, and diagnoses to standardized concepts. For medications, we first constructed a standardized drug concept database combining the nSights knowledge graph^57^ with RXNorm (https://www.nlm.nih.gov/research/umls/rxnorm/index.html) hierarchies to capture ingredient, brand, and dose-specific information. EHR medication records were matched using a hierarchical approach prioritizing RXNorm codes when available, followed by ingredient-level matching, and finally natural language processing and pattern matching on free-text medication orders when structured codes were absent. For anthropometric measurements (height, weight, BMI), we created a unified vocabulary from SNOMED (https://www.snomed.org/, https://athena.ohdsi.org) and LOINC (https://loinc.org/) terminologies and matched EHR measurement descriptions using standardized text matching algorithms with abbreviation expansion and synonym resolution; ambiguous mappings were resolved using OpenAI GPT-4o (https://platform.openai.com/docs/models/gpt-4o) with summary statistics as context, followed by manual verification. For diagnoses, we developed a hierarchical disease concept database from the nSights knowledge graph and matched EHR diagnosis descriptions and codes by identifying the most specific common child concept in the hierarchy. This approach enabled consistent identification of clinical entities while preserving granularity where available.

## Data Availability

This study involves the analysis of de-identified Electronic Health Record (EHR) data via the nference nSights Federated Clinical Analytics Platform (nSights). Data shown and reported in this manuscript were extracted from this environment using an established protocol for data extraction, aimed at preserving patient privacy. The data has been de-identified pursuant to an expert determination in accordance with the HIPAA Privacy Rule. Any data beyond what is reported in the manuscript, including but not limited to the raw EHR data, cannot be shared or released due to the parameters of the expert determination to maintain the data de-identification. Contact the corresponding author for additional details regarding nSights.

## Conflict of Interest Statement

The authors are employees of nference, inc., which conducts research collaborations with various biopharmaceutical companies, including AstraZeneca, Eli Lilly and Company, and Novo Nordisk A/S, whose GLP-1 receptor agonist products (exenatide, dulaglutide, tirzepatide, liraglutide, and semaglutide formulations) are included in this study. None of these companies, nor any other nference collaborator, funded, supported, or had any role in the independent study design, data acquisition, analysis, interpretation, manuscript preparation, or the decision to submit this work for publication. All analyses were conducted by the authors using de-identified electronic health record data. The authors declare no additional competing interests.

## Acknowledgements

We thank Pulkit Khandelwal, Prateek Singh, Harish Ashok Kumar, Vineet Agarwal, Ayush Mittal, Sankar Ardhanari, Viswanathan Thiagarajan, and the nference engineering team for development and operation of the nSights federated AI platform. We also thank Marcelo Montorzi and Praveen Kumar for their clinical guidance and interpretation, and Pradeep Rajendran and Christopher Gregg for their review of the manuscript.

## Supplementary Information

**Table S1.** Brand details for the GLP-1RAs included in this study.

| Brand | Drug | Approval Date | Min Dose (mg) | Max Dose (mg) | Route | Frequency |
| --- | --- | --- | --- | --- | --- | --- |
| Ozempic | Semaglutide | Dec 2017 | 0.25 | 2 | Subcutaneous | Weekly |
| Wegovy | Semaglutide | June 2021 | 0.25 | 2.4 | Subcutaneous | Weekly |
| Mounjaro | Tirzepatide | May 2022 | 2.5 | 15 | Subcutaneous | Weekly |
| Zepbound | Tirzepatide | Nov 2023 | 2.5 | 15 | Subcutaneous | Weekly |
| Saxenda | Liraglutide | Dec 2014 | 0.6 | 3 | Subcutaneous | Daily |
| Victoza | Liraglutide | Jan 2010 | 0.6 | 1.8 | Subcutaneous | Daily |
| Trulicity | Dulaglutide | Sept 2014 | 0.75 | 4.5 | Subcutaneous | Weekly |
| Byetta | Exenatide | Apr 2005 | 5 | 20 | Subcutaneous | Twice Daily |
| Bydureon | Exenatide | Jan 2012 | 0 | 2 | Subcutaneous | Weekly |
| Rybelsus | Semaglutide | Sept 2019 | 3 | 14 | Oral | Daily |

**Table S2.** Demographic characteristics of non–propensity-matched cohorts of weight-loss super responders, moderate responders, the minimal weight-loss group, and the weight regain group across GLP-1RA drugs and brands.

| DRUG<br>(Active Compound) | BRAND (Marketed Formulation) | Weight-Loss Outcome Category | Number of Patients (N) | Male (N) | Female (N) | Male Percentage (%) | Gender Association – log<sub>10</sub>(P) | Age in years, Mean (SD) | Age in years, Median [IQR] | Pregnant Patients N (%) | Race: White N (%) | Race: Black/African American N (%) | Race: Hispanic N (%) | Race: Other/Unknown N (%) |
| --- | --- | --- | --- | --- | --- | --- | --- | --- | --- | --- | --- | --- | --- | --- |
| All GLP-1 Receptor Agonists |  | Minimal weight-loss group | 63753 | 26207 | 37538 | 41.1 | inf | 54.58 (13.50) | 56 [45, 65] | 678 (1.1%) | 52582 (83.0%) | 5799 (9.1%) | 1366 (2.2%) | 3637 (5.7%) |
| All GLP-1 Receptor Agonists |  | Weight regain | 7365 | 2497 | 4867 | 33.9 | 167.17 | 52.09 (14.08) | 52 [42, 63] | 139 (1.9%) | 6362 (86.8%) | 521 (7.1%) | 119 (1.6%) | 331 (4.5%) |
| All GLP-1 Receptor Agonists |  | Moderate Responder | 47196 | 16817 | 30376 | 35.6 | inf | 55.28 (13.60) | 57 [46, 66] | 479 (1%) | 40447 (85.9%) | 3502 (7.4%) | 867 (1.8%) | 2257 (4.8%) |
| All GLP-1 Receptor Agonists |  | Super Responder | 16950 | 3375 | 13573 | 19.9 | inf | 51.52 (13.58) | 52 [41, 62] | 229 (1.4%) | 15247 (90.1%) | 867 (5.1%) | 178 (1.1%) | 637 (3.8%) |
| Semaglutide | (Ozempic / Wegovy / Rybelsus) | Minimal weight-loss group | 16122 | 7284 | 8836 | 45.2 | 33.63 | 56.15 (13.87) | 57 [47, 66] | 106 (1.7%) | 8190 (82.7%) | 756 (7.6%) | 369 (3.7%) | 589 (5.9%) |
| Semaglutide | (Ozempic / Wegovy / Rybelsus) | Weight regain | 1960 | 751 | 1208 | 38.3 | 24.27 | 53.86 (14.62) | 55 [43, 66] | 31 (1.6%) | 1072 (87.2%) | 73 (5.9%) | 16 (1.3%) | 68 (5.5%) |
| Semaglutide | (Ozempic / Wegovy / Rybelsus) | Moderate Responder | 14819 | 5731 | 9087 | 38.7 | 166.59 | 57.63 (13.73) | 59 [49, 68] | 77 (0.5%) | 8866 (86.8%) | 622 (6.1%) | 235 (2.3%) | 491 (4.8%) |
| Semaglutide | (Ozempic / Wegovy / Rybelsus) | Super Responder | 4601 | 889 | 3710 | 19.3 | inf | 53.47 (14.48) | 54 [43, 65] | 47 (1.0%) | 2907 (91.2%) | 125 (3.9%) | 27 (0.8%) | 129 (4.0%) |
| Tirzepatide | (Mounjaro / Zepbound) | Minimal weight-loss group | 4474 | 1684 | 2789 | 37.6 | 60.59 | 52.42 (13.69) | 53 [43, 63] | 55 (1.2%) | 2544 (80.4%) | 319 (10.1%) | 98 (3.1%) | 203 (6.4%) |
| Tirzepatide | (Mounjaro / Zepbound) | Weight regain | 794 | 210 | 584 | 26.4 | 39.48 | 48.81 (13.42) | 49 [39, 59] | 31 (3.9%) | 450 (86.7%) | 18 (3.5%) | 17 (3.3%) | 34 (6.6%) |
| Tirzepatide | (Mounjaro / Zepbound) | Moderate Responder | 7345 | 2731 | 4613 | 37.2 | 106.17 | 53.47 (13.43) | 54 [44, 64] | 61 (0.8%) | 4141 (84.5%) | 390 (8.0%) | 125 (2.5%) | 247 (5.0%) |
| Tirzepatide | (Mounjaro / Zepbound) | Super Responder | 5367 | 1164 | 4203 | 21.7 | inf | 50.65 (13.03) | 50 [41, 60] | 49 (0.9%) | 2811 (89.0%) | 155 (4.9%) | 54 (1.7%) | 137 (4.3%) |
| (Semaglutide) | **Ozempic** | Minimal weight-loss group | 8436 | 4405 | 4031 | 52.2 | 4.33 | 58.14 (12.98) | 60 [50, 67] | 50 (0.6%) | 6960 (82.5%) | 627 (7.4%) | 347 (4.1%) | 502 (5.9%) |
| (Semaglutide) | **Ozempic** | Weight regain | 1006 | 442 | 563 | 44 | 3.87 | 57.22 (13.35) | 59 [48, 67] | <11 (<1%) | 870 (87.3%) | 63 (6.3%) | 16 (1.6%) | 48 (4.8%) |
| (Semaglutide) | **Ozempic** | Moderate Responder | 8278 | 3586 | 4692 | 43.3 | 33.27 | 60.16 (12.54) | 62 [53, 69] | 31 (0.4%) | 7154 (86.4%) | 499 (6.0%) | 235 (2.8%) | 390 (4.7%) |
| (Semaglutide) | **Ozempic** | Super Responder | 1941 | 495 | 1446 | 25.5 | 102.61 | 59.11 (13.01) | 61 [50, 68] | <11 (<1%) | 1748 (90.6%) | 84 (4.4%) | 27 (1.4%) | 71 (3.7%) |
| (Tirzepatide) | **Mounjaro** | Minimal weight-loss group | 2410 | 1061 | 1348 | 44 | 8.3 | 55.06 (13.37) | 56 [46, 65] | 20 (0.8%) | 1230 (83.8%) | 129 (8.8%) | 22 (1.5%) | 87 (5.9%) |
| (Tirzepatide) | **Mounjaro** | Weight regain | 452 | 139 | 313 | 30.8 | 15.56 | 50.67 (13.46) | 51 [41, 60] | 10 (2.2%) | 202 (87.1%) | 10 (4.3%) | <11 (<1%) | 20 (8.6%) |
| (Tirzepatide) | **Mounjaro** | Moderate Responder | 3565 | 1535 | 2030 | 43.1 | 15.95 | 56.42 (12.83) | 57 [48, 66] | 20 (0.6%) | 1712 (88.4%) | 123 (6.4%) | <11 (<1%) | 101 (5.2%) |
| (Tirzepatide) | **Mounjaro** | Super Responder | 2039 | 524 | 1515 | 25.7 | 106.03 | 53.93 (12.94) | 55 [45, 64] | 16 (0.8%) | 1159 (92.1%) | 41 (3.3%) | <11 (<1%) | 58 (4.6%) |
| (Semaglutide) | **Wegovy** | Minimal weight-loss group | 1468 | 391 | 1076 | 26.7 | 70.81 | 48.16 (14.37) | 48 [38, 58] | 15 (1%) | 1928 (80.0%) | 249 (10.3%) | 83 (3.4%) | 150 (6.2%) |
| (Semaglutide) | **Wegovy** | Weight regain | 239 | 60 | 179 | 25.1 | 13.86 | 46.62 (14.04) | 45 [36, 58] | 10 (4.3%) | 388 (85.8%) | 13 (2.9%) | 17 (3.8%) | 34 (7.5%) |
| (Semaglutide) | **Wegovy** | Moderate Responder | 1936 | 546 | 1389 | 28.2 | 81.13 | 49.81 (14.33) | 50 [40, 60] | 22 (1.1%) | 3009 (84.4%) | 273 (7.7%) | 102 (2.9%) | 181 (5.1%) |
| (Semaglutide) | **Wegovy** | Super Responder | 1258 | 179 | 1077 | 14.3 | 140.92 | 46.93 (13.53) | 47 [37, 56] | 23 (1.8%) | 1814 (89.0%) | 107 (5.2%) | 37 (1.8%) | 81 (4.0%) |
| (Tirzepatide) | **Zepbound** | Minimal weight-loss group | 754 | 209 | 545 | 27.7 | 33.7 | 47.82 (13.67) | 48 [37, 58] | 19 (2.5%) | 616 (81.7%) | 70 (9.3%) | 15 (2.0%) | 53 (7.0%) |
| (Tirzepatide) | **Zepbound** | Weight regain | 67 | <11 | N/A | <17 | 12.25 | 45.42 (14.14) | 45 [34, 57] | <11 (<16%) | 62 (92.5%) | 5 (7.5%) | <11 (<1%) | 0 (0.0%) |
| (Tirzepatide) | **Zepbound** | Moderate Responder | 1338 | 407 | 931 | 30.4 | 45.82 | 48.33 (13.02) | 48 [39, 57] | 21 (1.6%) | 1132 (84.6%) | 117 (8.7%) | 23 (1.7%) | 66 (5.0%) |
| (Tirzepatide) | **Zepbound** | Super Responder | 1118 | 196 | 922 | 17.5 | 103.81 | 46.04 (12.36) | 47 [37, 55] | 13 (1.2%) | 997 (89.2%) | 48 (4.3%) | 17 (1.5%) | 56 (5.1%) |

**Table S3.**
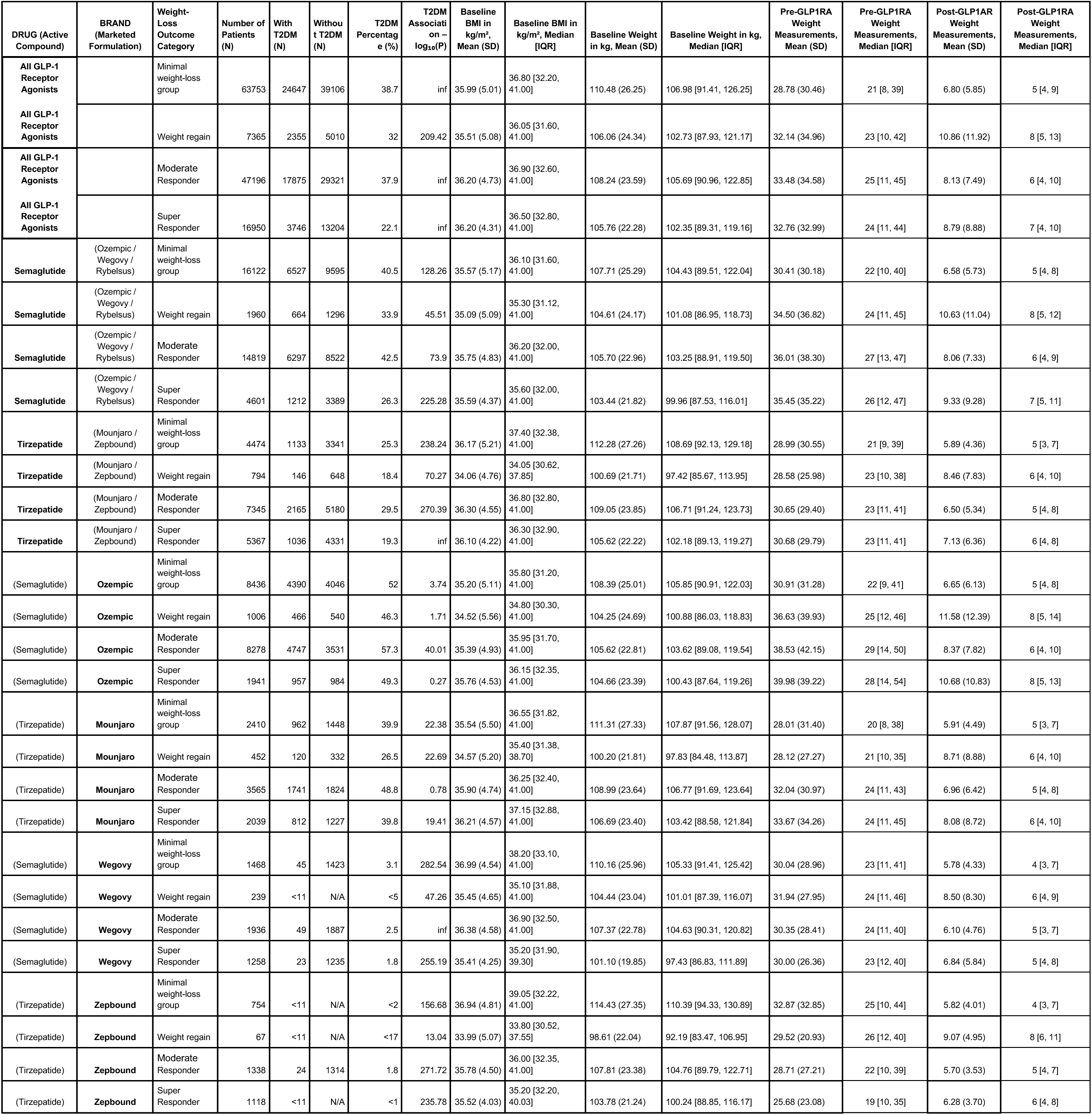
Baseline metabolic conditions and longitudinal weight and BMI measurements before and after GLP-1RA therapy in non-propensity-matched cohorts of weight-loss super responders, moderate responders, the minimal weight-loss group, and the weight regain group across GLP-1RA drugs and brands.

**Table S4.** Duration and number of prescriptions, number of clinical documents per patient per month among non-propensity-matched cohorts of weight loss super responders, moderate responders, minimal weight-loss group, and weight regain.

| DRUG (Active Compound) | BRAND (Marketed Formulation) | Weight-Loss Outcome Category | Number of Patients (N) | Duration of prescriptions per patient in months, Mean (SD) | Duration of prescriptions per patient in months, Median [IQR] | Number of prescriptions per patient, Mean (SD) | Number of prescriptions per patient, Median [IQR] | Baseline monthly document per patient, Mean (SD) | Baseline monthly documents per patient, Median [IQR] | Post-treatment monthly documents per patient, Mean (SD) | Post-treatment monthly documents per patient, Median [IQR] |
| --- | --- | --- | --- | --- | --- | --- | --- | --- | --- | --- | --- |
| All GLP-1 Receptor Agonists |  | Minimal weight-loss group | 63753 | 27.90 (26.84) | 21.28 [10.62, 36.59] | 13.24 (11.50) | 10 [6, 18] | 7.6 (16.9) | 3 [1, 7] | 6.2 (11.7) | 3 [2, 7] |
| All GLP-1 Receptor Agonists |  | Weight regain | 7365 | 25.19 (22.76) | 20.60 [11.60, 31.95] | 14.89 (13.00) | 11 [6, 20] | 8.3 (19.7) | 3 [2, 8] | 8.8 (22.8) | 4 [2, 8] |
| All GLP-1 Receptor Agonists |  | Moderate Responder | 47196 | 22.08 (20.30) | 17.65 [8.99, 28.72] | 15.02 (11.85) | 11 [7, 20] | 7.3 (16.7) | 3 [1, 7] | 6.2 (12.9) | 3 [2, 6] |
| All GLP-1 Receptor Agonists |  | Super Responder | 16950 | 18.23 (13.51) | 15.54 [9.16, 24.65] | 17.73 (13.04) | 15 [9, 23] | 6.7 (16.4) | 3 [1, 6] | 6.2 (15.9) | 3 [2, 6] |
| Semaglutide | (Ozempic / Wegovy / Rybelsus) | Minimal weight-loss group | 16122 | 17.16 (13.08) | 14.15 [6.71, 24.01] | 9.77 (7.47) | 8 [4, 13] | 7.2 (15.8) | 3 [1, 7] | 5.8 (10.9) | 3 [2, 6] |
| Semaglutide | (Ozempic / Wegovy / Rybelsus) | Weight regain | 1960 | 16.84 (12.37) | 14.56 [7.16, 23.14] | 10.91 (8.23) | 9 [6, 15] | 7.9 (18.5) | 3 [2, 7] | 8.4 (20.2) | 4 [2, 7] |
| Semaglutide | (Ozempic / Wegovy / Rybelsus) | Moderate Responder | 14819 | 16.76 (12.11) | 13.90 [7.29, 23.33] | 11.51 (8.48) | 9 [6, 15] | 6.9 (16.1) | 3 [2, 6] | 6.0 (12.0) | 3 [2, 6] |
| Semaglutide | (Ozempic / Wegovy / Rybelsus) | Super Responder | 4601 | 16.76 (10.93) | 14.74 [8.50, 22.91] | 13.54 (9.57) | 11 [6, 18] | 6.7 (18.5) | 3 [2, 6] | 6.5 (15.8) | 3 [2, 6] |
| Tirzepatide | (Mounjaro / Zepbound) | Minimal weight-loss group | 4474 | 11.10 (7.95) | 8.90 [4.70, 15.95] | 10.69 (8.50) | 8 [5, 14] | 7.2 (15.2) | 3 [1, 7] | 5.1 (7.7) | 3 [1, 6] |
| Tirzepatide | (Mounjaro / Zepbound) | Weight regain | 794 | 14.44 (8.90) | 13.27 [7.11, 21.46] | 14.85 (11.13) | 12 [6, 20] | 6.8 (12.9) | 3 [1, 7] | 5.6 (11.7) | 3 [1, 6] |
| Tirzepatide | (Mounjaro / Zepbound) | Moderate Responder | 7345 | 11.70 (7.73) | 9.76 [5.81, 16.57] | 14.29 (10.34) | 12 [7, 19] | 6.2 (13.5) | 3 [1, 6] | 4.9 (8.2) | 3 [1, 6] |
| Tirzepatide | (Mounjaro / Zepbound) | Super Responder | 5367 | 13.33 (7.80) | 11.57 [7.59, 18.42] | 17.58 (12.29) | 15 [10, 23] | 6.0 (13.3) | 3 [1, 6] | 4.7 (8.7) | 3 [1, 6] |
| (Semaglutide) | Ozempic | Minimal weight-loss group | 8436 | 16.98 (13.23) | 13.71 [6.72, 23.75] | 9.46 (7.04) | 8 [4, 11] | 7.7 (16.8) | 3 [2, 7] | 5.8 (11.3) | 3 [2, 7] |
| (Semaglutide) | Ozempic | Weight regain | 1006 | 17.16 (12.75) | 14.30 [7.83, 23.52] | 10.00 (7.90) | 8 [5, 13] | 8.8 (20.1) | 3 [2, 8] | 9.1 (20.7) | 4 [2, 8] |
| (Semaglutide) | Ozempic | Moderate Responder | 8278 | 17.03 (12.45) | 14.00 [7.36, 23.81] | 10.65 (7.62) | 9 [6, 14] | 7.7 (17.2) | 3 [2, 7] | 6.3 (12.0) | 3 [2, 6] |
| (Semaglutide) | Ozempic | Super Responder | 1941 | 17.20 (12.02) | 14.55 [8.07, 24.10] | 11.39 (8.04) | 9 [6, 15] | 8.2 (23.4) | 4 [2, 7] | 8.1 (19.4) | 4 [2, 7] |
| (Tirzepatide) | Mounjaro | Minimal weight-loss group | 2410 | 10.84 (7.85) | 8.49 [4.50, 15.66] | 10.25 (7.93) | 8 [5, 14] | 7.4 (16.0) | 3 [1, 7] | 5.1 (8.0) | 3 [1, 6] |
| (Tirzepatide) | Mounjaro | Weight regain | 452 | 12.34 (8.25) | 10.25 [6.25, 17.05] | 12.91 (9.17) | 11 [6, 17] | 7.1 (14.0) | 3 [1, 7] | 6.3 (14.5) | 3 [1, 6] |
| (Tirzepatide) | Mounjaro | Moderate Responder | 3565 | 12.11 (7.82) | 10.15 [5.95, 17.35] | 13.36 (9.60) | 11 [7, 17] | 6.9 (15.4) | 3 [1, 6] | 5.3 (9.3) | 3 [2, 6] |
| (Tirzepatide) | Mounjaro | Super Responder | 2039 | 14.27 (8.33) | 12.12 [7.77, 20.72] | 16.61 (11.81) | 14 [8, 22] | 7.2 (15.9) | 3 [1, 6] | 5.6 (10.2) | 3 [2, 6] |
| (Semaglutide) | Wegovy | Minimal weight-loss group | 1468 | 9.98 (7.81) | 7.56 [3.91, 14.30] | 8.04 (5.57) | 6 [5, 10] | 5.7 (12.5) | 3 [2, 6] | 4.5 (7.7) | 3 [1, 5] |
| (Semaglutide) | Wegovy | Weight regain | 239 | 10.81 (7.70) | 8.83 [5.17, 14.75] | 11.03 (8.21) | 9 [6, 13] | 4.6 (10.2) | 2 [1, 5] | 4.6 (9.2) | 2 [1, 5] |
| (Semaglutide) | Wegovy | Moderate Responder | 1936 | 11.04 (7.40) | 9.04 [5.44, 14.98] | 10.96 (7.05) | 9 [6, 14] | 4.0 (8.2) | 2 [1, 4] | 3.6 (7.8) | 2 [1, 3] |
| (Semaglutide) | Wegovy | Super Responder | 1258 | 13.62 (7.78) | 12.24 [7.96, 17.82] | 13.46 (8.33) | 11 [8, 17] | 3.9 (8.1) | 2 [1, 4] | 3.5 (7.3) | 2 [1, 3] |
| (Tirzepatide) | Zepbound | Minimal weight-loss group | 754 | 6.71 (4.12) | 5.82 [3.43, 9.53] | 8.51 (6.32) | 6 [4, 11] | 6.9 (14.8) | 3 [2, 7] | 5.4 (7.8) | 3 [2, 6] |
| (Tirzepatide) | Zepbound | Weight regain | 67 | 8.09 (4.76) | 8.22 [4.01, 11.73] | 12.21 (9.41) | 9 [6, 15] | 5.2 (8.4) | 3 [1, 5] | 4.3 (4.6) | 3 [2, 5] |
| (Tirzepatide) | Zepbound | Moderate Responder | 1338 | 7.22 (3.91) | 6.70 [4.19, 9.71] | 12.25 (8.42) | 10 [6, 16] | 5.5 (11.4) | 3 [1, 6] | 4.3 (5.7) | 3 [1, 5] |
| (Tirzepatide) | Zepbound | Super Responder | 1118 | 8.90 (3.75) | 8.94 [5.94, 11.70] | 15.27 (10.01) | 13 [8, 20] | 4.9 (8.8) | 3 [1, 5] | 4.1 (5.4) | 3 [1, 5] |

**Table S5.** Treatment-emergent adverse events among GLP-1RA super responders with FDR-corrected p<0.001. Post-treatment disease prevalence increases among super responders across four GLP-1 receptor agonist formulations. Rate ratios represent pre-treatment prevalence divided by post-treatment prevalence, with values <1.0 indicating increased post-treatment occurrence (adverse events). Of the 1,426 disease terms analyzed, 36 unique terms were identified with FDR-corrected p<0.001 and rate ratio <=0.5. Final columns categorize adverse events as brand-specific, differentially severe between brands, or shared across all (or >=3) brands. Rows are shaded as Ozempic-specific signals (red), Mounjaro-specific signals (orange), Zepbound-specific signals (purple), Wegovy-specific signals (yellow), and common signals across brands (blue).

| Brand | Clinical Variable | Category | Rate Ratio | FDR Corrected<br>Negative Log P Value | Only significant<br>adverse event in | More adverse events<br>in | All (or $\geq 3$ drugs) have<br>adverse events |
| --- | --- | --- | --- | --- | --- | --- | --- |
| ozempic | acidosis | diseases | 0.18 | 3.3 | Ozempic |  |  |
| ozempic | acute blood loss anemia | diseases | 0.29 | 3.28 | Ozempic |  |  |
| mounjaro | acute cystitis | diseases | 0.5 | 4.36 |  | Ozempic more AEs<br>than Mounjaro |  |
| ozempic | acute cystitis | diseases | 0.37 | 9.99 |  | Ozempic more AEs<br>than Mounjaro |  |
| ozempic | acute kidney injury | diseases | 0.41 | 6.79 |  | Mounjaro more AEs<br>than Ozempic |  |
| mounjaro | acute kidney injury | diseases | 0.3 | 5.64 |  | Mounjaro more AEs<br>than Ozempic |  |
| mounjaro | alopecia | diseases | 0.26 | 3.52 | Mounjaro |  |  |
| ozempic | bacteremia | diseases | 0.17 | 3.47 | Ozempic |  |  |
| ozempic | bilateral pleural effusion | textdiseases | 0.14 | 4.61 | Ozempic |  |  |
| mounjaro | constipation | diseases | 0.39 | 7.45 |  |  | All |
| ozempic | constipation | diseases | 0.33 | 11.79 |  |  | All |
| wegovy | constipation | diseases | 0.33 | 6.75 |  |  | All |
| zepbound | constipation | diseases | 0.27 | 4.36 |  |  | All |
| ozempic | decreased appetite | textdiseases | 0.32 | 3.22 | Ozempic |  |  |
| mounjaro | dehydration | diseases | 0.21 | 5.12 |  | Ozempic more AEs<br>than Mounjaro |  |
| ozempic | dehydration | diseases | 0.19 | 10.13 |  | Ozempic more AEs<br>than Mounjaro |  |
| mounjaro | dizziness and giddiness symptoms | diseases | 0.44 | 7.4 | Mounjaro |  |  |
| mounjaro | generalized abdominal pain | diseases | 0.27 | 3.76 | Mounjaro |  |  |
| ozempic | hematuria | diseases | 0.46 | 5.27 | Ozempic |  |  |
| ozempic | hypercalcemia | diseases | 0.21 | 5.51 | Ozempic |  |  |
| mounjaro | hypokalemia | diseases | 0.37 | 5.93 |  | Zepbound > Ozempic ><br>Mounjaro |  |
| ozempic | hypokalemia | diseases | 0.34 | 9.65 |  | Zepbound > Ozempic ><br>Mounjaro |  |
| zepbound | hypokalemia | diseases | 0.18 | 3.31 |  | Zepbound > Ozempic ><br>Mounjaro |  |
| ozempic | hypomagnesemia | diseases | 0.38 | 3.17 | Ozempic |  |  |
| ozempic | hyponatremia | diseases | 0.48 | 3.22 |  | Mounjaro more AEs<br>than Ozempic |  |
| mounjaro | hyponatremia | diseases | 0.19 | 6.24 |  | Mounjaro more AEs<br>than Ozempic |  |
| ozempic | hypotension | diseases | 0.23 | 8.06 |  | Ozempic and Mounjaro<br>have this AE |  |
| mounjaro | hypotension | diseases | 0.09 | 8.88 |  | Ozempic and Mounjaro<br>have this AE |  |
| ozempic | hypotensive | textdiseases | 0.12 | 5.99 |  | Ozempic and Mounjaro<br>have this AE |  |
| mounjaro | insulin resistance | diseases | 0.35 | 3.47 | Mounjaro |  |  |
| mounjaro | iron deficiency | diseases | 0.33 | 3.12 | Mounjaro |  |  |
| ozempic | malnutrition | textdiseases | 0.11 | 6.61 | Ozempic |  |  |
| mounjaro | nausea | diseases | 0.43 | 6.34 |  |  | All |
| ozempic | nausea | diseases | 0.42 | 8.08 |  |  | All |
| wegovy | nausea | diseases | 0.38 | 5.98 |  |  | All |
| zepbound | nausea | diseases | 0.35 | 6.01 |  |  | All |
| wegovy | nausea and vomiting | diseases | 0.37 | 5.19 | Wegovy |  |  |
| ozempic | nausea and vomiting | diseases | 0.24 | 13.34 | Ozempic |  |  |
| mounjaro | orthostatic hypotension | diseases | 0.19 | 3.12 |  | Ozempic more AEs<br>than Mounjaro |  |
| ozempic | orthostatic hypotension | diseases | 0.1 | 7.64 |  | Ozempic more AEs<br>than Mounjaro |  |
| ozempic | pleural effusion | diseases | 0.17 | 3.66 | Ozempic |  |  |
| ozempic | protein energy malnutrition | diseases | 0.24 | 7.49 |  | Mounjaro more AEs<br>than Ozempic |  |
| mounjaro | protein energy malnutrition | diseases | 0.17 | 3.47 |  | Mounjaro more AEs<br>than Ozempic |  |
| zepbound | pure hypercholesterolemia | diseases | 0.45 | 3.36 | Zepbound |  |  |
| ozempic | sepsis | diseases | 0.28 | 4.38 | Ozempic |  |  |
| mounjaro | swollen lymph nodes | diseases | 0.3 | 3.32 | Mounjaro |  |  |
| mounjaro | syncope | diseases | 0.31 | 4.56 |  | Mounjaro == Ozempic |  |
| ozempic | syncope | diseases | 0.31 | 6.4 |  | Mounjaro == Ozempic |  |
| ozempic | urinary retention | diseases | 0.19 | 3.12 | Ozempic |  |  |
| ozempic | urinary tract infection | diseases | 0.46 | 5.22 | Ozempic |  |  |
| ozempic | vomiting | diseases | 0.09 | 8.88 | Ozempic |  |  |
| ozempic | weakness | diseases | 0.39 | 7.92 | Ozempic |  |  |

**Table S6.** Demographic characteristics of age– and gender–propensity-matched cohorts of weight-loss super responders and the minimal weight-loss group across GLP-1RA drugs and brands.

| DRUG<br>(Active Compound) | BRAND<br>(Marketed Formulation) | Weight-Loss Outcome Category | Number of Patients (N) | Male (N) | Female (N) | Male Percent age (%) | Gender Association $-\log_{10}(P)$ | Age in years, Mean ± SD (years) | Age, Median [IQR] (years) | Pregnant Patients N (%) | Race: White N (%) | Race: Black/African American N (%) | Race: Hispanic N (%) | Race: Other/Unknown N (%) |
| --- | --- | --- | --- | --- | --- | --- | --- | --- | --- | --- | --- | --- | --- | --- |
| All GLP-1RA |  | Minimal weight-loss group | 16948 | 3376 | 13572 | 19.9 | inf | 51.52 (13.57) | 52 [41, 62] | 209 (1.2%) | 14022 (82.7%) | 1474 (8.7%) | 541 (3.2%) | 1009 (5.9%) |
| All GLP-1RA |  | Super Responder | 16948 | 3375 | 13573 | 19.9 | inf | 51.52 (13.58) | 52 [41, 62] | 157 (0.9%) | 15230 (89.9%) | 749 (4.4%) | 282 (1.7%) | 737 (4.3%) |
| Semaglutide | (Ozempic / Wegovy / Rybelsus) | Minimal weight-loss group | 4599 | 889 | 3710 | 19.3 | inf | 53.48 (14.49) | 54 [43, 65] | 46 (1.0%) | 3905 (84.9%) | 344 (7.5%) | 162 (3.5%) | 242 (5.3%) |
| Semaglutide | (Ozempic / Wegovy / Rybelsus) | Super Responder | 4599 | 889 | 3710 | 19.3 | inf | 53.48 (14.48) | 54 [43, 65] | 41 (0.9%) | 4182 (90.9%) | 185 (4.0%) | 64 (1.4%) | 189 (4.1%) |
| Tirzepatide | (Mounjaro / Zepbound) | Minimal weight-loss group | 3857 | 1068 | 2789 | 27.7 | 168.29 | 51.99 (13.65) | 53 [42, 62] | 58 (1.5%) | 3096 (80.3%) | 388 (10.1%) | 113 (2.9%) | 259 (6.7%) |
| Tirzepatide | (Mounjaro / Zepbound) | Super Responder | 3857 | 1024 | 2833 | 26.5 | 185.8 | 51.66 (12.67) | 52 [43, 61] | 37 (1.0%) | 3420 (88.7%) | 188 (4.9%) | 72 (1.9%) | 178 (4.6%) |
| (Semaglutide) | **Ozempic** | Minimal weight-loss group | 1941 | 495 | 1446 | 25.5 | 102.61 | 59.11 (13.01) | 61 [50, 68] | 14 (0.7%) | 1633 (84.6%) | 137 (7.1%) | 68 (3.5%) | 92 (4.8%) |
| (Semaglutide) | **Ozempic** | Super Responder | 1941 | 495 | 1446 | 25.5 | 102.61 | 59.11 (13.01) | 61 [50, 68] | <11 (<1%) | 1748 (90.6%) | 84 (4.4%) | 27 (1.4%) | 71 (3.7%) |
| (Tirzepatide) | **Mounjaro** | Minimal weight-loss group | 1866 | 545 | 1321 | 29.2 | 71.43 | 54.59 (13.25) | 55 [45, 64] | 20 (1.1%) | 839 (85.5%) | 81 (8.3%) | N/A | 61 (6.2%) |
| (Tirzepatide) | **Mounjaro** | Super Responder | 1866 | 524 | 1342 | 28.1 | 79.24 | 53.93 (12.83) | 54 [45, 64] | 15 (0.8%) | 899 (91.6%) | 33 (3.4%) | N/A | 49 (5.0%) |
| (Semaglutide) | **Wegovy** | Minimal weight-loss group | 981 | 179 | 802 | 18.2 | 87.31 | 47.58 (14.20) | 48 [37, 57] | 15 (1.5%) | 1490 (79.8%) | 194 (10.4%) | 61 (3.3%) | 121 (6.5%) |
| (Semaglutide) | **Wegovy** | Super Responder | 981 | 179 | 802 | 18.2 | 87.31 | 47.26 (13.12) | 47 [38, 56] | 16 (1.6%) | 1658 (88.9%) | 99 (5.3%) | 34 (1.8%) | 75 (4.0%) |
| (Tirzepatide) | **Zepbound** | Minimal weight-loss group | 719 | 174 | 545 | 24.2 | 42.81 | 47.11 (13.44) | 47 [37, 57] | 19 (2.6%) | 585 (81.4%) | 66 (9.2%) | 15 (2.1%) | 53 (7.4%) |
| (Tirzepatide) | **Zepbound** | Super Responder | 719 | 154 | 565 | 21.4 | 52.3 | 47.03 (12.10) | 47 [38, 56] | <11 (<2%) | 634 (88.2%) | 27 (3.8%) | 14 (1.9%) | 44 (6.1%) |

**Table S7.**
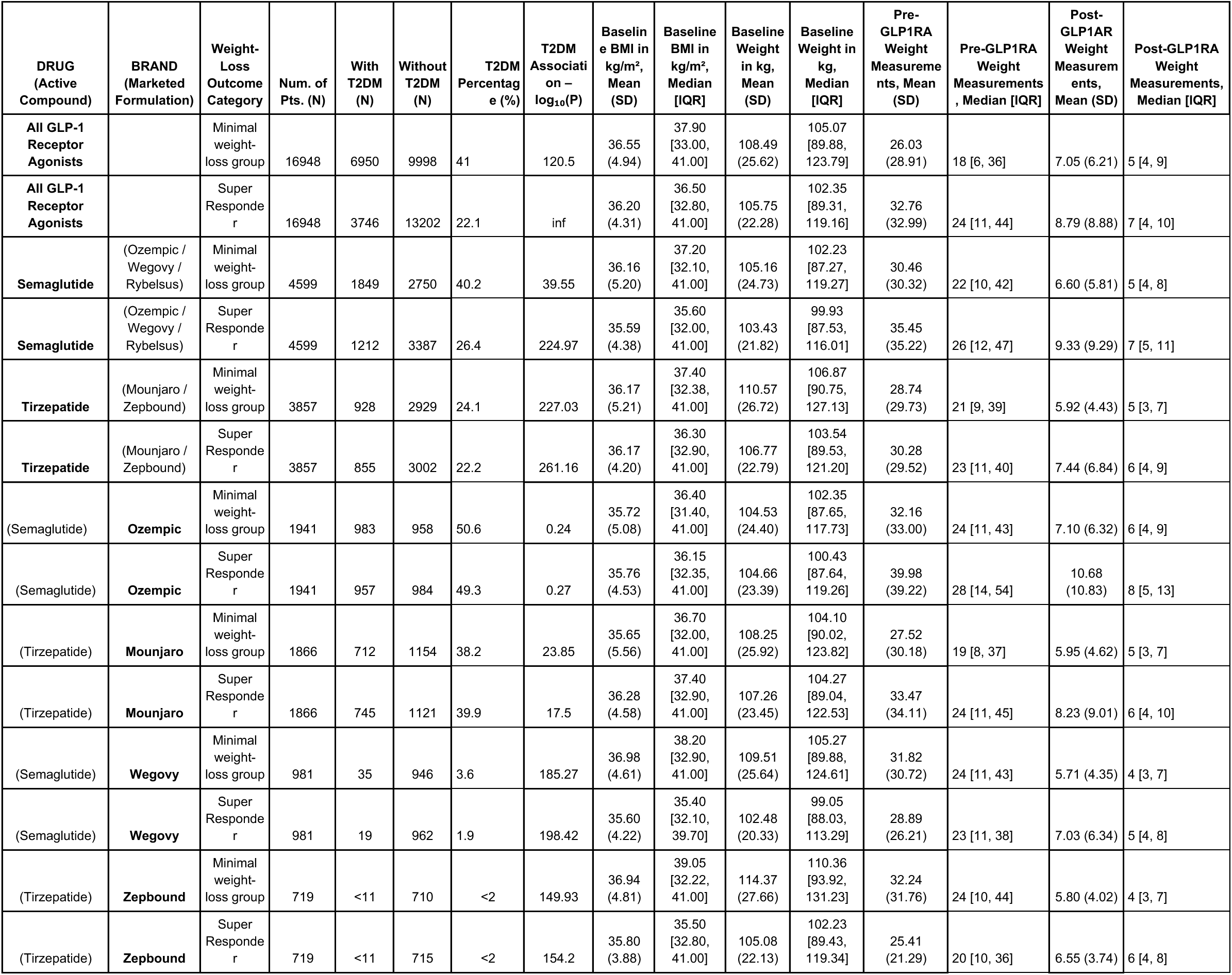
Baseline metabolic conditions and longitudinal weight and BMI measurements before and after GLP-1RA therapy in age– and gender–propensity-matched cohorts of weight-loss super responders and minimal weight-loss group.

**Table S8.**
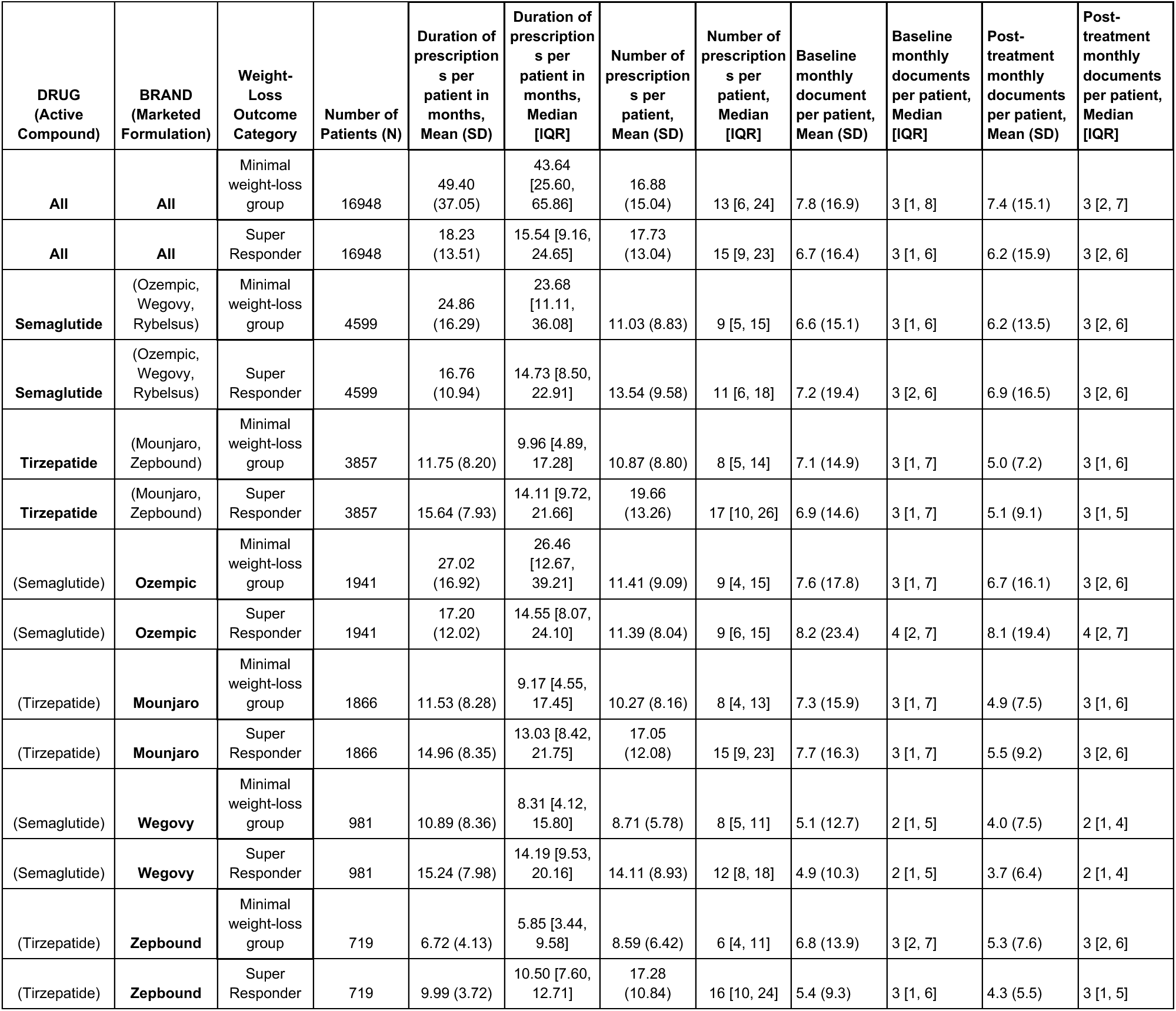
Duration and number of prescriptions, number of clinical documents per patient per month among age– and gender–propensity-matched cohorts of weight-loss super responders and minimal weight-loss group.

| DRUG<br>(Active Compound) | BRAND<br>(Marketed Formulation) | Weight-Loss Outcome Category | Number of Patients (N) | Duration of prescriptions per patient in months, Mean (SD) | Duration of prescriptions per patient in months, Median [IQR] | Number of prescriptions per patient, Mean (SD) | Number of prescriptions per patient, Median [IQR] | Baseline monthly document per patient, Mean (SD) | Baseline monthly documents per patient, Median [IQR] | Post-treatment monthly documents per patient, Mean (SD) | Post-treatment monthly documents per patient, Median [IQR] |
| --- | --- | --- | --- | --- | --- | --- | --- | --- | --- | --- | --- |
| All | All | Minimal weight-loss group | 16948 | 49.40 (37.05) | 43.64 [25.60, 65.86] | 16.88 (15.04) | 13 [6, 24] | 7.8 (16.9) | 3 [1, 8] | 7.4 (15.1) | 3 [2, 7] |
| All | All | Super Responder | 16948 | 18.23 (13.51) | 15.54 [9.16, 24.65] | 17.73 (13.04) | 15 [9, 23] | 6.7 (16.4) | 3 [1, 6] | 6.2 (15.9) | 3 [2, 6] |
| Semaglutide | (Ozempic, Wegovy, Rybelsus) | Minimal weight-loss group | 4599 | 24.86 (16.29) | 23.68 [11.11, 36.08] | 11.03 (8.83) | 9 [5, 15] | 6.6 (15.1) | 3 [1, 6] | 6.2 (13.5) | 3 [2, 6] |
| Semaglutide | (Ozempic, Wegovy, Rybelsus) | Super Responder | 4599 | 16.76 (10.94) | 14.73 [8.50, 22.91] | 13.54 (9.58) | 11 [6, 18] | 7.2 (19.4) | 3 [2, 6] | 6.9 (16.5) | 3 [2, 6] |
| Tirzepatide | (Mounjaro, Zepbound) | Minimal weight-loss group | 3857 | 11.75 (8.20) | 9.96 [4.89, 17.28] | 10.87 (8.80) | 8 [5, 14] | 7.1 (14.9) | 3 [1, 7] | 5.0 (7.2) | 3 [1, 6] |
| Tirzepatide | (Mounjaro, Zepbound) | Super Responder | 3857 | 15.64 (7.93) | 14.11 [9.72, 21.66] | 19.66 (13.26) | 17 [10, 26] | 6.9 (14.6) | 3 [1, 7] | 5.1 (9.1) | 3 [1, 5] |
| (Semaglutide) | Ozempic | Minimal weight-loss group | 1941 | 27.02 (16.92) | 26.46 [12.67, 39.21] | 11.41 (9.09) | 9 [4, 15] | 7.6 (17.8) | 3 [1, 7] | 6.7 (16.1) | 3 [2, 6] |
| (Semaglutide) | Ozempic | Super Responder | 1941 | 17.20 (12.02) | 14.55 [8.07, 24.10] | 11.39 (8.04) | 9 [6, 15] | 8.2 (23.4) | 4 [2, 7] | 8.1 (19.4) | 4 [2, 7] |
| (Tirzepatide) | Mounjaro | Minimal weight-loss group | 1866 | 11.53 (8.28) | 9.17 [4.55, 17.45] | 10.27 (8.16) | 8 [4, 13] | 7.3 (15.9) | 3 [1, 7] | 4.9 (7.5) | 3 [1, 6] |
| (Tirzepatide) | Mounjaro | Super Responder | 1866 | 14.96 (8.35) | 13.03 [8.42, 21.75] | 17.05 (12.08) | 15 [9, 23] | 7.7 (16.3) | 3 [1, 7] | 5.5 (9.2) | 3 [2, 6] |
| (Semaglutide) | Wegovy | Minimal weight-loss group | 981 | 10.89 (8.36) | 8.31 [4.12, 15.80] | 8.71 (5.78) | 8 [5, 11] | 5.1 (12.7) | 2 [1, 5] | 4.0 (7.5) | 2 [1, 4] |
| (Semaglutide) | Wegovy | Super Responder | 981 | 15.24 (7.98) | 14.19 [9.53, 20.16] | 14.11 (8.93) | 12 [8, 18] | 4.9 (10.3) | 2 [1, 5] | 3.7 (6.4) | 2 [1, 4] |
| (Tirzepatide) | Zepbound | Minimal weight-loss group | 719 | 6.72 (4.13) | 5.85 [3.44, 9.58] | 8.59 (6.42) | 6 [4, 11] | 6.8 (13.9) | 3 [2, 7] | 5.3 (7.6) | 3 [2, 6] |
| (Tirzepatide) | Zepbound | Super Responder | 719 | 9.99 (3.72) | 10.50 [7.60, 12.71] | 17.28 (10.84) | 16 [10, 24] | 5.4 (9.3) | 3 [1, 6] | 4.3 (5.5) | 3 [1, 5] |

**Figure S1.**
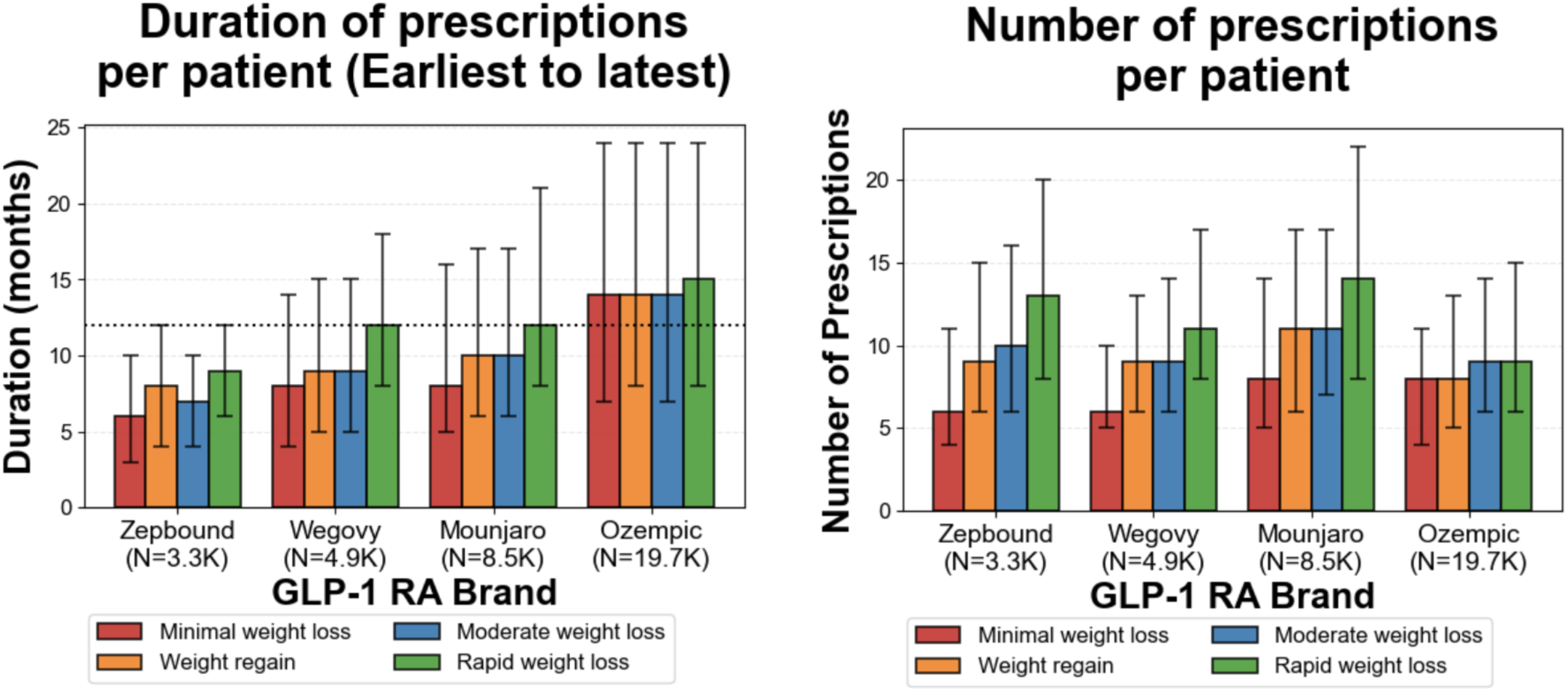
The four categories examined are the minimal weight loss group (red), weight regain group (orange), moderate responders (moderate weight loss; blue) and super responders (rapid weight loss). (Left) Median duration of GLP-1RA brand-level prescriptions per patient for each response category (first prescription – index date, to last prescription even if beyond the observation period of 1 year post first prescription). Dotted line indicates 12-month threshold. **(Right)** Median number of GLP-1RA brand-level prescriptions per patient for each response category. In both plots, error bars represent interquartile range (IQR).

**Figure S2.**
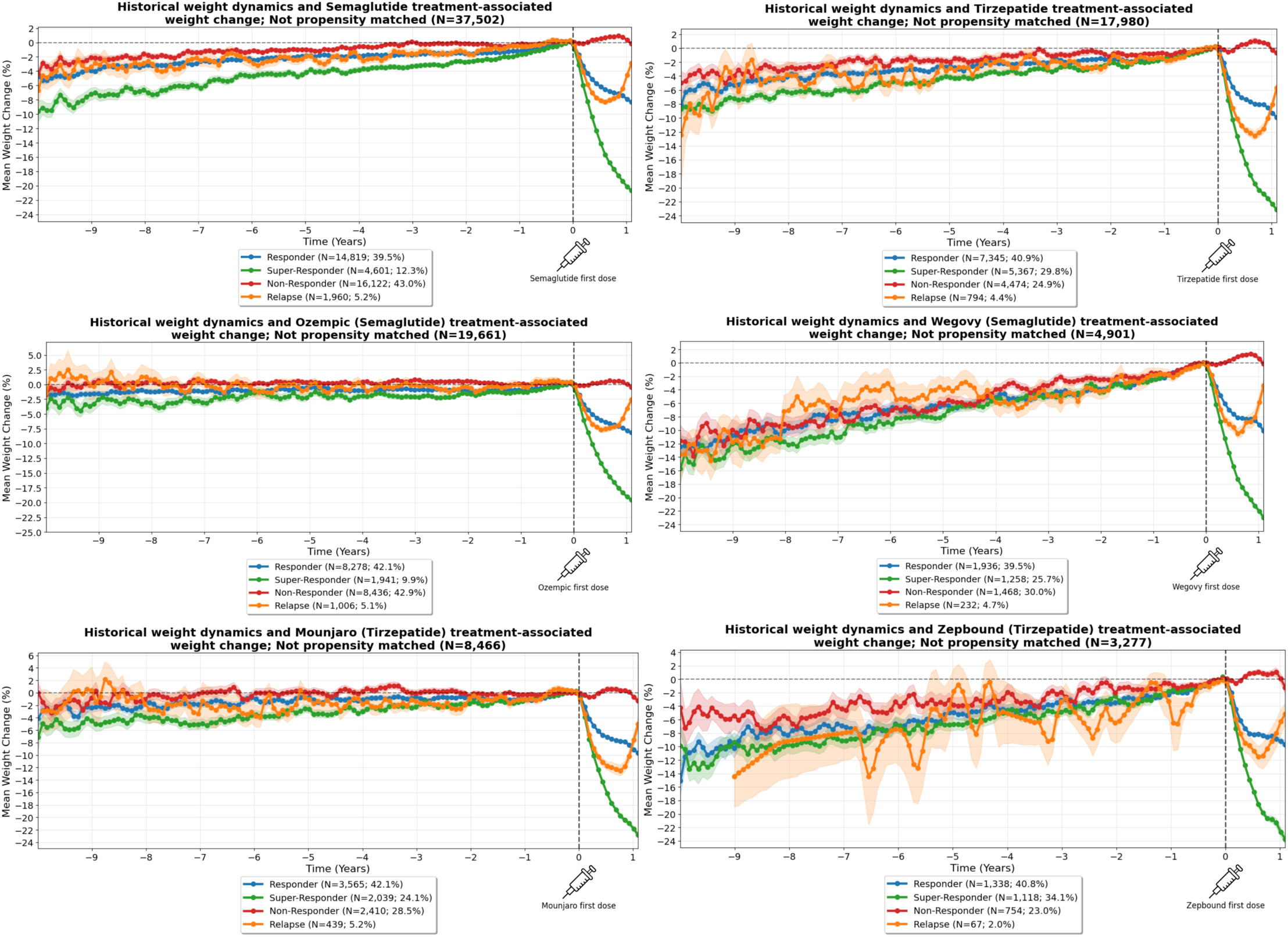
GLP-1RA brand specific 10-year historical weight dynamics and weight loss patterns upon GLP-1RA therapy initiation. All four response categories are shown: Super responder (green), moderate responder (blue), minimal weight-loss group (“Non-Responder”; red) and weight regain group (“Relapse”; orange).

**Figure S3.**
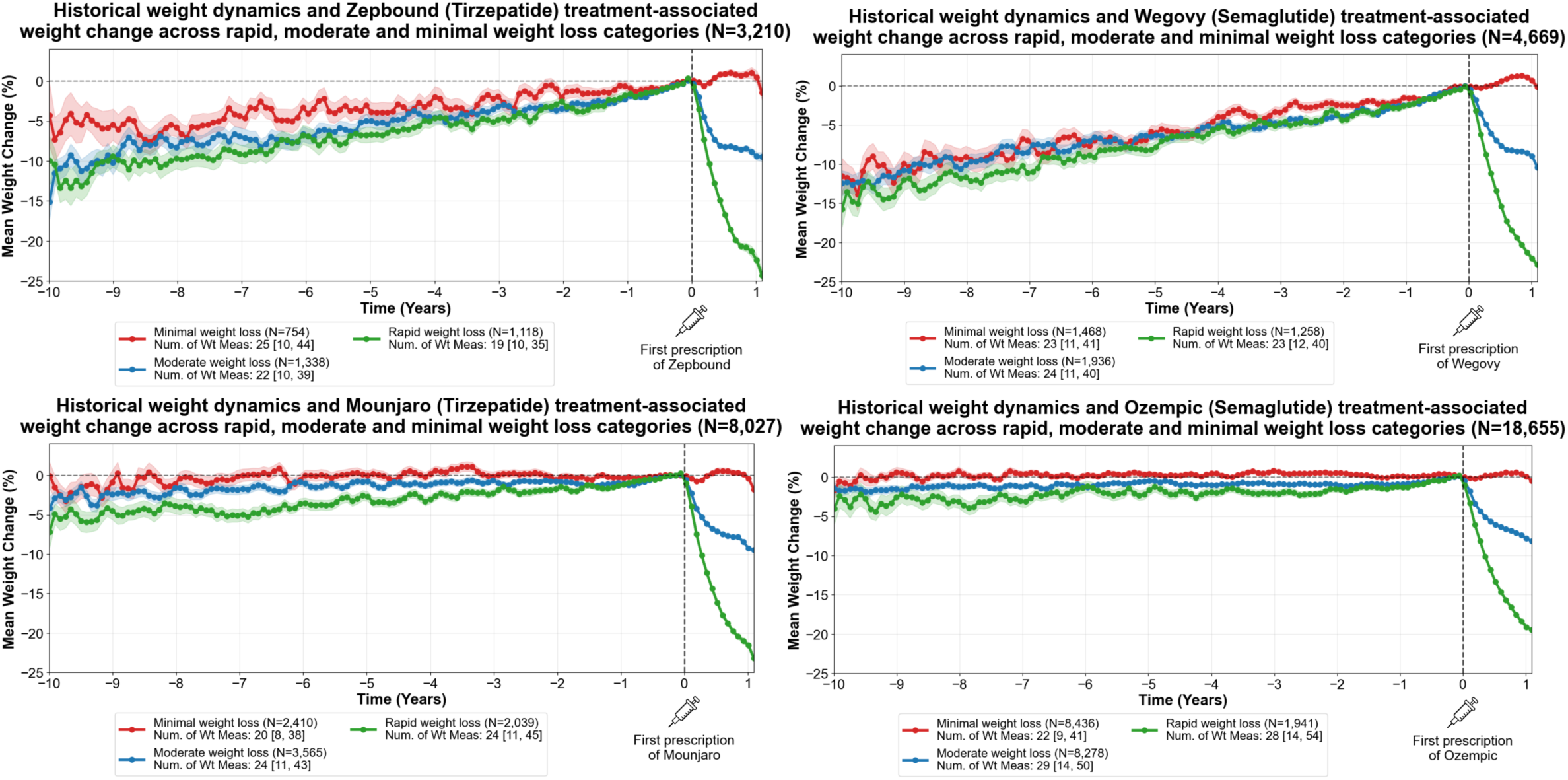
GLP-1RA brand specific 10-year historical weight dynamics and weight loss patterns upon GLP-1RA therapy initiation. Three response categories are shown: Super responder (green), moderate responder (blue), and the minimal weight-loss group (“Non-Responder”; red).

**Figure S4.**
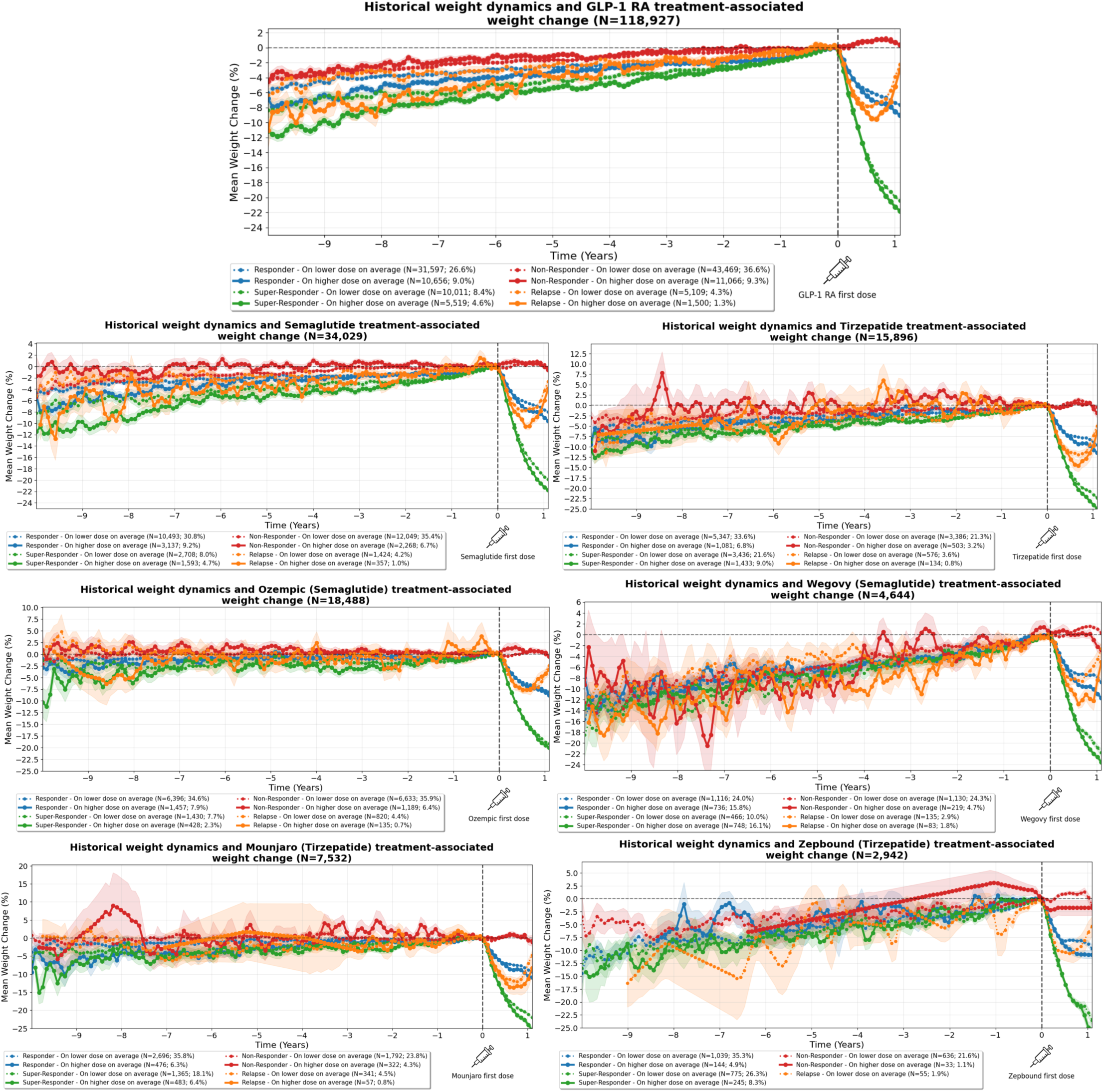
Historical weight loss trajectories for 10-years preceding GLP-1RA therapy initiation and high-dose/low-dose average prescriptions-associated weight loss trajectories post GLP-1RA initiation. All 4 response categories are shown: Super responder (green), moderate responder (blue), minimal weight-loss group (“Non-Responder”; red) and weight regain group (“Relapse”; orange).

## Supplementary Text (Figure S4 Methodology)

To enable comparison across GLP-1RAs formulations with different therapeutic dose ranges, each prescription was normalized to a 0-1 scale using the formula:

Normalized Dose = (Actual Dose – Minimum Therapeutic Dose) / (Maximum Therapeutic Dose – Minimum Therapeutic Dose), where minimum and maximum doses were defined based on FDA-approved therapeutic ranges for each brand (Table S1).

A normalized dose of 0 represents the minimum starting dose (e.g., 0.25 mg for semaglutide, 2.5 mg for tirzepatide), while 1.0 represents the maximum target dose (e.g., 2.4 mg for Wegovy, 15 mg for Mounjaro). For each patient, a time-weighted average dose proportion was calculated across all prescriptions during the treatment period, with each prescription’s normalized dose weighted by its duration of use (defined as the time interval until the next prescription, or 30 days for the final prescription):

Average Dose Proportion = Σ(Normalized Dose × Duration) / Σ(Duration).

Patients were then stratified into dose cohorts based on their average dose proportion: ‘lower dose on average’ (<0.5, below the therapeutic range midpoint) and ‘higher dose on average’ (>=0.5, at or above the therapeutic range midpoint), and weight-loss trajectories were compared between dose cohorts (Figure S4). Only prescriptions with valid dose units (mg) and dose amounts within the established therapeutic range for each brand were included in dose calculations.

## Supplementary Text (Figure S4 Interpretation)

Dose-stratified analyses revealed divergence in post-treatment weight loss trajectories between higher and lower dose cohorts across all response categories. The observed dose-dependent effects suggest that dose optimization strategies may represent an important area for future investigation. Further research examining dosing patterns, dose escalation protocols, and their impact on long-term weight outcomes could help establish evidence-based guidelines for maximizing personalized therapeutic benefit from GLP-1RA therapy.

**Figure S5:**
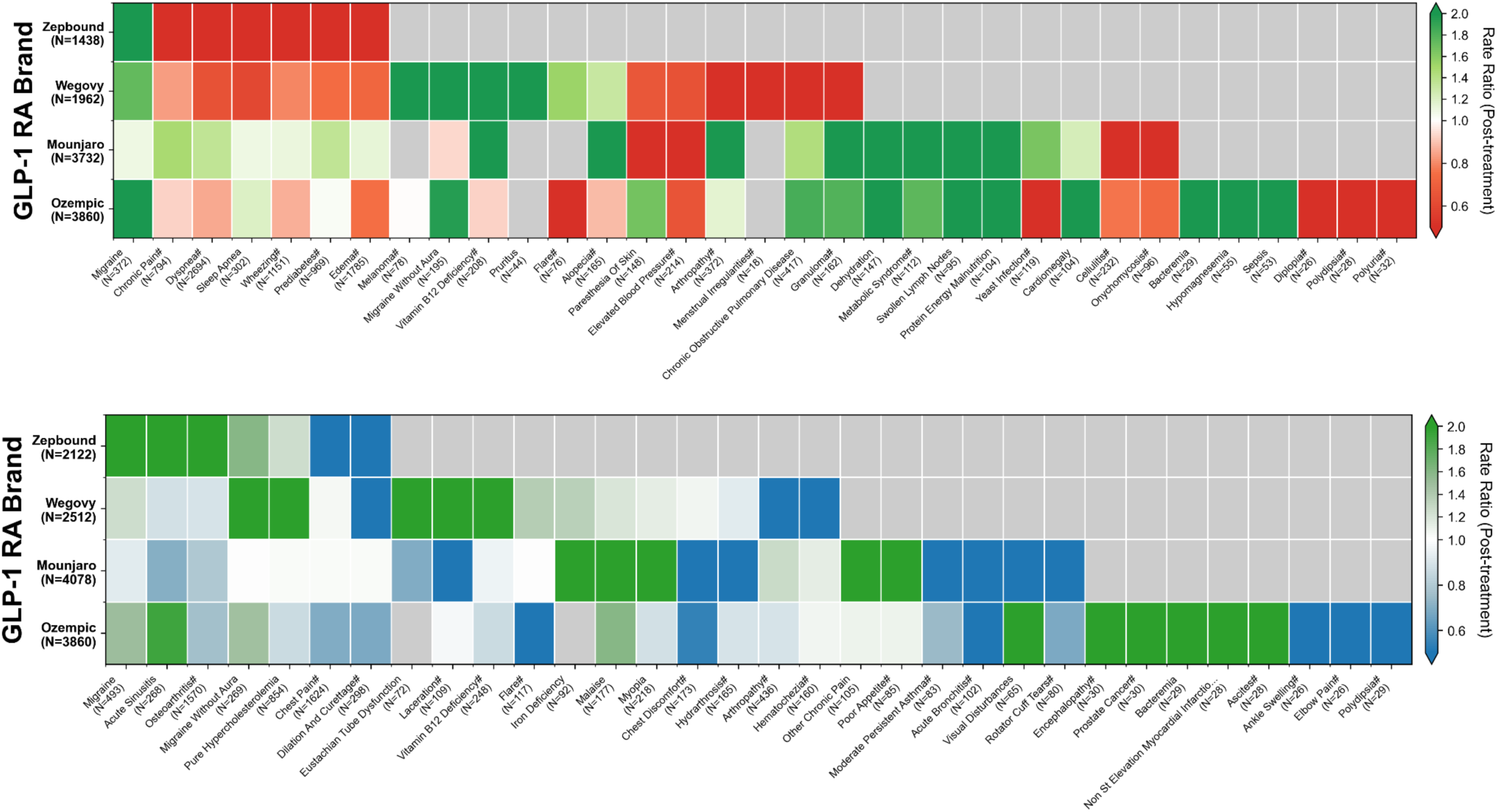
*(**Top**)* The hallmark phenotypes of super responders versus the minimal weight-loss group during the first year of initiating each GLP-1RA treatment colored by the Rate Ratio (post-treatment initiation prevalence for super responders/minimal weight-loss group). *(**Bottom**)* The hallmark phenotypes of super responders versus moderate responders during the first year of initiating each GLP-1RA treatment colored by the Rate Ratio (post-treatment initiation prevalence for super responders/moderate responders).

## Supplementary Text (Figure S5 Interpretation)

### Zepbound super responders

The super responder cohort exhibited elevated post-treatment incidence relative to the minimal weight-loss cohort for migraine (RR=2.4, p=0.024; Figure S5, top). In contrast, super responders experienced substantially lower post-treatment incidence of chronic pain (RR=0.22, p=0.004), dyspnea (RR=0.22, p<0.001), sleep apnea (RR=0.17, p<0.001), wheezing (RR=0.16, p<0.001), prediabetes (RR=0.12, p<0.001) and edema (RR=0.12, p<0.001). Super responders relative to moderate responders displayed higher incidence of migraine (RR=3.0, p=0.002), acute sinusitis (RR=3.0, p=0.016), and osteoarthritis (RR=2.1, p=0.014; Figure S5, bottom).

### Wegovy super responders

In the post-treatment period following Wegovy therapy, the super responder cohort exhibited relative enrichment over the minimal weight-loss cohort for melanoma (RR=5.2, p=0.001), migraine without aura (RR=3.1, p=0.003), vitamin b12 deficiency (RR=2.5, p=0.012), and pruritus (RR=2.1, p=0.049; Figure S5, top). Conversely, arthropathy (RR=0.3, p=0.001), menstrual irregularities (RR=0.3, p=0.026), chronic obstructive pulmonary disease (RR=0.2, p=0.002), and granuloma (RR=0.2, p<0.001) were depleted in the post-treatment period for the super responder cohort relative to the minimal weight-loss cohort. When evaluating differences between super responders and moderate responders, post-treatment enrichments were noted in the super responder cohort for eustachian tube dysfunction (RR=5.2, p=0.003), laceration (RR=4.0, p=0.025), migraine without aura (RR=3.1, p=0.005), pure hypercholesterolemia (RR=3.0, p=0.004), and vitamin b12 deficiency (RR=2.3, p=0.019), while depletion of incidence in the rapid loss cohort was seen for dilation and curettage (RR=0.5, p=0.028), arthropathy (RR=0.5, p=0.031), and hematochezia (RR=0.4, p=0.026; Figure S5, bottom).

### Mounjaro super responders

Following Mounjaro therapy, the super responder cohort demonstrated higher relative incidence over the minimal weight-loss cohort for hypotension (RR=10.6, p<0.001), dehydration (RR=9.4, p<0.001), metabolic syndrome (RR=9.2, p<0.001), alopecia (RR=8.8, p<0.001), swollen lymph nodes (RR=8.0, p<0.001), protein energy malnutrition (RR=5.6, p<0.001), mediastinal disorders (RR=5.6, p<0.001), arthropathy (RR=2.5, p<0.001), and vitamin b12 deficiency (RR=2.4, p=0.011), whereas lower incidence was observed in the super responder cohort for paresthesia of skin (RR=0.4, p=0.029), elevated blood pressure (RR=0.4, p=0.029), cellulitis (RR=0.4, p=0.006), and onychomycosis (RR=0.2, p=0.008; Figure S5, top). Examining the distinction between super responders and moderate responders, enrichments in the super responder cohort were noted for iron deficiency (RR=9.0, p<0.001), myopia (RR=8.8, p<0.001), other chronic pain (RR=6.4, p<0.001), poor appetite (RR=6.2, p<0.001), and malaise (RR=6.2, p<0.001), while laceration (RR=0.3, p=0.025), acute bronchitis (RR=0.2, p=0.013), moderate persistent asthma (RR=0.2, p=0.013), rotator cuff tears (RR=0.2, p=0.003), visual disturbances (RR=0.2, p=0.002), chest discomfort (RR=0.2, p=0.001), and hydrarthrosis (RR=0.2, p<0.001) were enriched in moderate responders (Figure S5, bottom).

### Ozempic super responders

Among patients prescribed Ozempic, post-treatment enrichments in super responders relative to minimal weight-loss patients included bacteremia (RR>15, p<0.001), hypotension (RR=15.8, p<0.001), protein energy malnutrition (RR=15.2, p<0.001), hypomagnesemia (RR=11.0, p<0.001), cardiomegaly (RR=10.6, p<0.001), and sepsis (RR=10.6, p<0.001). The minimal weight-loss cohort showed higher post-treatment incidence of yeast infection (RR=0.2, p<0.001), diplopia (RR=0.2, p=0.001), polydipsia (RR=0.2, p<0.001), flare (RR=0.2, p<0.001), and polyuria (RR=0.2, p<0.001; Figure S5, top). When comparing super responders versus moderate responders, the former cohort exhibited higher relative incidence of encephalopathy (RR=6.0, p<0.001), prostate cancer (RR=6.0, p<0.001), bacteremia (RR=5.8, p<0.001), craniocerebral trauma (RR=5.6, p=0.001), Non–ST-elevation myocardial infarction (NSTEMI) (RR=5.6, p=0.001), and ascites (RR=5.6, p=0.001), while moderate responders had enrichment for chest discomfort (RR=0.5, p=0.017), ankle swelling (RR=0.2, p=0.003), elbow pain (RR=0.2, p=0.003), polydipsia (RR=0.2, p<0.001), and flare (RR=0.2, p<0.001; Figure S5, bottom).

**Figure S6.**
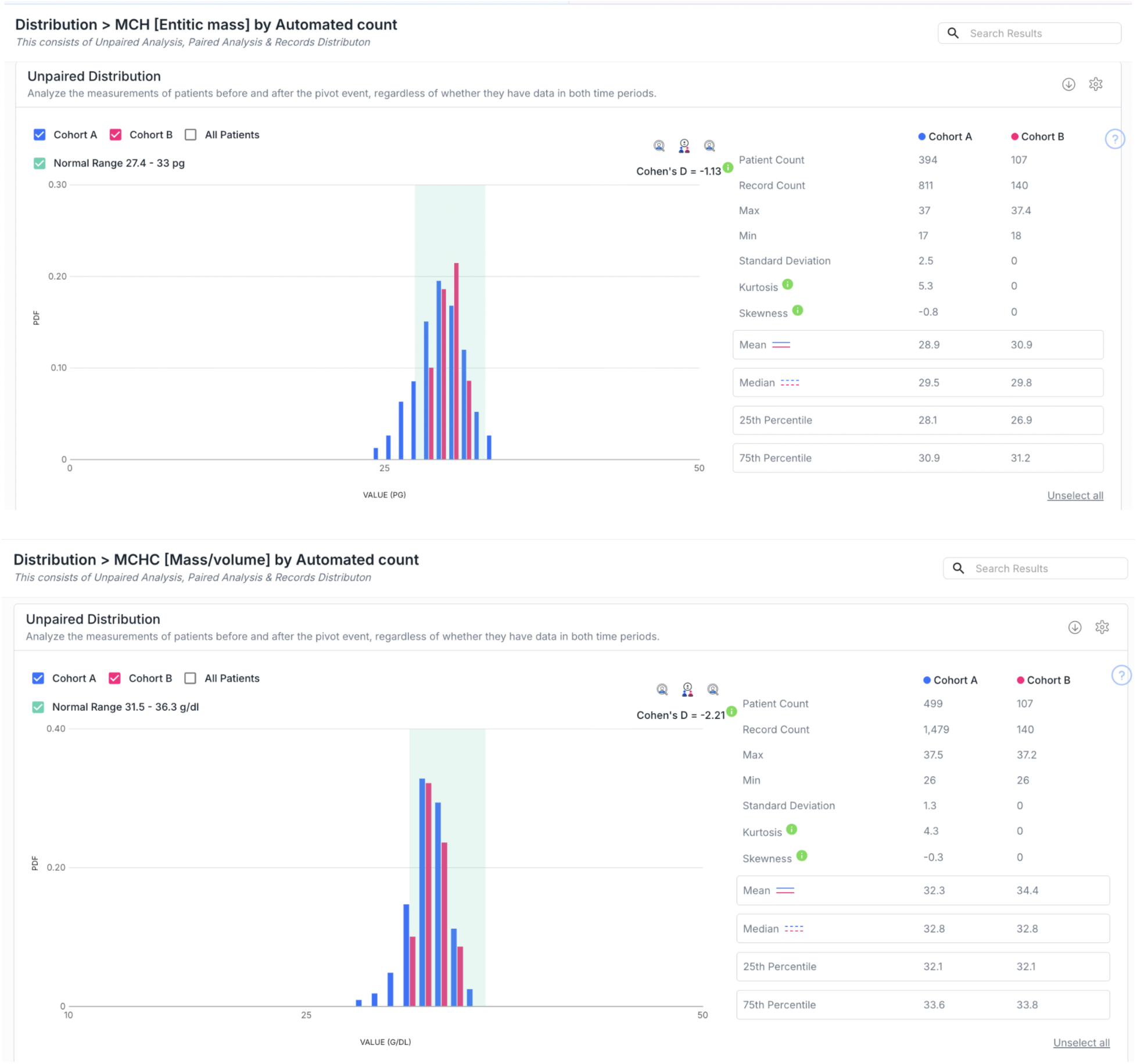
nSights platform shows significant effect size separation (Cohen’s D) for – MCH [Entitic Mass] *(top)* and MCHC [Mass/Volume] lab test (*bottom)* distributions – for Zepbound (tirzepatide) minimal weight-loss group (N=754) during the 1-year post first Zepbound prescription (pink; Cohort B) relative to the same minimal-weight loss patient population in the 1-year prior to that index date of first Zepbound prescription (blue, Cohort A). For both MCH and MCHC, the post-treatment distributions in the Zepbound minimal weight-loss group shift noticeably toward the center of the established normal range, with a larger proportion of values falling within physiologic intervals compared with the pre-treatment period. This pattern indicates that tirzepatide can subtly normalize or recalibrate red-cell indices even in patients who do not exhibit meaningful weight loss, suggesting that hematologic effects may occur independently of clinical response. Although values largely remain within reference limits, the systematic pre-to-post movement toward the normal band underscores the importance of longitudinal laboratory surveillance across all GLP-1RA cohorts, including individuals in the minimal weight-loss group, to detect evolving metabolic or nutritional changes that may not be apparent from weight trajectories alone.

## Notes

### Funding Statement

This study did not receive any external funding. All authors are employees of nference, inc., and no author or institution received payment or services from any third party for any aspect of the submitted work.

### Author Declarations

This study analyzed de-identified EHR data from a network of tertiary clinical centers tied to academic medical centers in the United States through the nference nSights Analytics Platform [55-56]. nference, in collaboration with academic medical center (AMC) data partners provided the de-identified data for this study. nference has established a secure data environment, hosted by and within each of the AMCs, that house the AMC's de-identified patient data. The provisioning of and access to this data are governed by an expert determination that satisfies the HIPAA Privacy Rule requirements for the de-identification of protected health information. Each AMC's de-identified data environment is specifically designed and operated to enable access to and analysis of de-identified data without the need for Institutional Review Board (IRB) oversight, approval, or an exemption confirmation. Given these measures, informed consent and IRB review were not required for this study. References: 55. Murugadoss, K. et al. Building a best-in-class automated de-identification tool for electronic health records through ensemble learning. Patterns (N Y) 2, 100255 (2021). 56. Murugadoss, K. et al. Scaling text de-identification using locally augmented ensembles. medRxiv 2024.06.20.24308896 (2024) doi:10.1101/2024.06.20.24308896.

